# Protection of second booster vaccinations and prior infection against SARS-CoV-2 in the UK SIREN healthcare worker cohort

**DOI:** 10.1101/2023.09.29.23296330

**Authors:** Peter D Kirwan, Victoria Hall, Sarah Foulkes, Ashley Otter, Katie Munro, Dominic Sparkes, Anna Howells, Naomi Platt, Jonathan Broad, David Crossman, Chris Norman, Diane Corrigan, Christopher H Jackson, Michelle Cole, Colin S Brown, Ana Atti, Jasmin Islam, SIREN Study Group, Anne M Presanis, Andre Charlett, Daniela De Angelis, Susan Hopkins

## Abstract

**Background:** The protection of fourth dose mRNA vaccination against SARS-CoV-2 is relevant to current global policy decisions regarding ongoing booster roll-out. We estimate the effect of fourth dose vaccination, prior infection, and duration of PCR positivity in a highly-vaccinated and largely prior-COVID-19 infected cohort of UK healthcare workers.

**Methods:** Participants underwent fortnightly PCR and regular antibody testing for SARS-CoV-2 and completed symptoms questionnaires. A multi-state model was used to estimate vaccine effectiveness (VE) against infection from a fourth dose compared to a waned third dose, with protection from prior infection and duration of PCR positivity jointly estimated.

**Results:** 1,298 infections were detected among 9,560 individuals under active follow-up between September 2022 and March 2023. Compared to a waned third dose, fourth dose VE was 13.1% (95%CI 0.9 to 23.8) overall; 24.0% (95%CI 8.5 to 36.8) in the first two months post-vaccination, reducing to 10.3% (95%CI - 11.4 to 27.8) and 1.7% (95%CI -17.0 to 17.4) at 2-4 and 4-6 months, respectively. Relative to an infection >2 years ago and controlling for vaccination, 63.6% (95%CI 46.9 to 75.0) and 29.1% (95%CI 3.8 to 43.1) greater protection against infection was estimated for an infection within the past 0-6, and 6-12 months, respectively. A fourth dose was associated with greater protection against asymptomatic infection than symptomatic infection, whilst prior infection independently provided more protection against symptomatic infection, particularly if the infection had occurred within the previous 6 months. Duration of PCR positivity was significantly lower for asymptomatic compared to symptomatic infection.

**Conclusions:** Despite rapid waning of protection, vaccine boosters remain an important tool in responding to the dynamic COVID-19 landscape; boosting population immunity in advance of periods of anticipated pressure, such as surging infection rates or emerging variants of concern.

**Funding:** UK Health Security Agency, Medical Research Council, NIHR HPRU Oxford, and others.

## Main text

### Background

More than three years since SARS-CoV-2 emerged, after 770 million reported infections and 13.5 billion doses of COVID-19 vaccine delivered [1], most of the world’s population now has some degree of immunity, whether from infection, vaccination or a hybrid of both. How long this protection lasts, against what outcome, and how future-proof it may be to emerging variants remains uncertain. With the public health emergency declared over by the World Health Organisation [2], budgets, testing, and vaccination programmes have been scaled back in many countries [3]. In the northern hemisphere, decisions about autumn vaccination campaigns are being made, with concomitant authorisation of vaccines which target emerging strains of SARS-CoV-2 [4].

In many high-income countries, following evidence of vaccine protection waning after the second dose [5–7], priority population groups have now been offered at least two ‘booster’ vaccinations (with up to 6 vaccines administered to the highest risk individuals) [8]. Healthcare workers without additional risk factors were offered their fourth vaccine dose in autumn 2022. This occurred during a period of dominance of Omicron sub-variants, which had higher immune escape capability from both vaccines and previous infection [9,10]. Together with the contemporaneous removal of non-pharmaceutical interventions [11] – and the return to more normal population mixing [12] – record levels of infections were detected [13]. Importantly, given a combination of high population immunity and variant properties, the Omicron sub-variant waves have been clinically milder than earlier variants [14]. Considerations about future vaccination campaigns must, therefore, be made in a context of population immunity, uncertainty about variant emergence, and the requirement for sustainable post-pandemic management of COVID-19. Healthcare workers have been prioritised for COVID-19 vaccination in the UK and many other countries, recognising their high occupational exposure, their potential role in nosocomial transmission dynamics, and the significant impact of healthcare worker absence on healthcare delivery [15]. Whether healthcare workers should be regularly offered autumn boosters is unresolved. There are opportunities for efficient operational delivery aligned with seasonal influenza vaccination, however, consideration of perceptions and behaviours affecting vaccine uptake, and the potential risk of vaccination fatigue, is important. Efficient timing of these vaccination campaigns should also be considered, for optimal protection during periods of highest COVID-19 and other respiratory virus circulation.

The SARS-CoV-2 Immunity and Reinfection Evaluation (SIREN) study, a large-scale United Kingdom (UK) healthcare worker cohort undergoing fortnightly PCR testing and running continuously since June 2020 [16], is uniquely placed to provide evidence to support this decision-making. Here we investigate the protection of booster vaccination and prior SARS-CoV-2 infection on the acquisition of infection (both symptomatic and asymptomatic) among our cohort of triple-vaccinated healthcare workers in a period of Omicron sub-variant circulation [5].

## Methods

### Study design and setting

The SIREN study, run by the UK Health Security Agency (UKHSA), is a prospective cohort study of National Health Service (NHS) healthcare workers, recruiting 44,000 participants between June 2020 and March 2021. Follow-up was initially 12 months from enrolment, with subsequent extensions to 24 and 36 months. At enrolment, participants completed a questionnaire and provided blood serum and nasal swab samples for anti-SARS-CoV-2 antibody testing and polymerase-chain reaction (PCR) testing for SARS-CoV-2 RNA. Following enrolment, participants provided samples for fortnightly PCR testing and regular antibody testing, and completed a fortnightly symptom questionnaire [16].

### Participant data

We included all participants maintained under active follow up between 12^th^ September 2022 and 31^st^ March 2023, with more than six months since receipt of their third dose and contributing at least two SARS-CoV-2 PCR tests during the follow-up period. We excluded participants who had received more than three doses before September 2022. Questionnaire responses, PCR and antibody test results (including from outside the study), and information on vaccination were collected centrally by UKHSA [16].

### Covariates

Demographic covariates included: age, gender, ethnicity, region of residence, occupational setting, staff type, medical conditions, and household structure (Table S3).

Linked testing and vaccination data included: PCR and antibody dates and results, vaccination dose, date, and manufacturer (with most receiving mRNA vaccines).

Participants were grouped by time since previous infection, where the date of a positive PCR test or a positive antibody test consistent with previous infection was used as a proxy for the onset of infection. For those without indication of prior infection we required an anti-N negative result (Roche Elecsys anti-SARS-CoV-2 nucleocapsid (anti-N) assay) within the 6 months prior to study entry to confirm their naïve status. Participants with indication of prior infection but missing serological information within the last 6 months were excluded from analyses exploring time since infection.

Questionnaires completed within a 14-day window of a positive PCR test were used to distinguish between infections with and without COVID-19 symptoms. We assigned symptom statuses of: COVID-19-specific symptoms (any of: cough, fever, sore throat, anosmia, and/or dysgeusia), and asymptomatic for COVID-19 (absence of symptoms, or only non-specific symptoms such as fatigue and muscle ache).

### Representativeness

All NHS hospitals and health boards in the UK were invited to join SIREN, with no random sampling of hospitals, health boards, or participants [16]. The cohort, whilst not representative of the general UK population, broadly reflects the demography of healthcare workers.

The Spikevax bivalent Original/Omicron and Comirnaty bivalent Original/Omicron booster vaccines received regulatory approval in August and September 2022, respectively, and were introduced for frontline healthcare workers on 12^th^ September 2022 (with eligibility criteria consistent with third dose criteria).

### Bias

The fortnightly testing regime minimised bias in detection of SARS-CoV-2 infection, and the statistical methodology further controlled for gaps in testing. Recall bias was minimised by only considering symptoms reported within a 14-day window of a positive PCR test.

### Censoring

Testing dates were pre-determined upon study enrolment, providing interval-censored observations of infection. Participants joined the study analysis at the date of their first PCR test after 12^th^ September 2022. Participants with fewer than 24 weeks since receiving a third vaccine dose were left-censored and only entered the study once sufficient time had elapsed. To account for study withdrawal (either because of early withdrawal or reaching the end of the follow-up period) participants were right-censored at their last recorded PCR test.

### Statistical methods

Crude PCR positivity rates were calculated as the number of detected PCR positive results per 10,000 person-days of follow-up. An exact Poisson method was used to calculate 95% confidence intervals (CI). Multi-state models (MSMs) were used to estimate hazards associated with infection for selected covariates and time spent in the PCR positive state.

We compared MSM estimates to a Cox proportional hazards model, assessed model fit by comparing expected and observed numbers in each state over time, and undertook variable selection using Akaike information criterion values, and likelihood-ratio tests. Stratification and piecewise-constant hazards over time were used to account for non-proportionality. Vaccine effectiveness (VE) and relative protection estimates were obtained from hazard ratios (HR) using the formula: VE = 1 – HR [7]. We used 2+ years as the baseline for time since previous infection because the confirmed naïve group was smaller and testing records indicated different behavioural trends, see Supplementary Appendix for further details.

Duration of positivity estimates were averaged over the demographic characteristics of the entire study population for comparability.

### Model implementation

Statistical models were implemented using R v.4.3.1 (R Foundation, Vienna, Austria) and the R package msm was used to fit the multi-state models.

## Results

### Population characteristics

A total of 9,560 participants were included in this analysis; 6,776 (70.9%) had a previous SARS-CoV-2 infection date recorded, and 773 (8.1%) were confirmed as naïve (Figure 1). Coverage of booster vaccination was low compared with previous vaccines; 64.3% uptake within two months of fourth dose availability, compared to 83.0% within two months of third dose availability.

**Figure 1:**
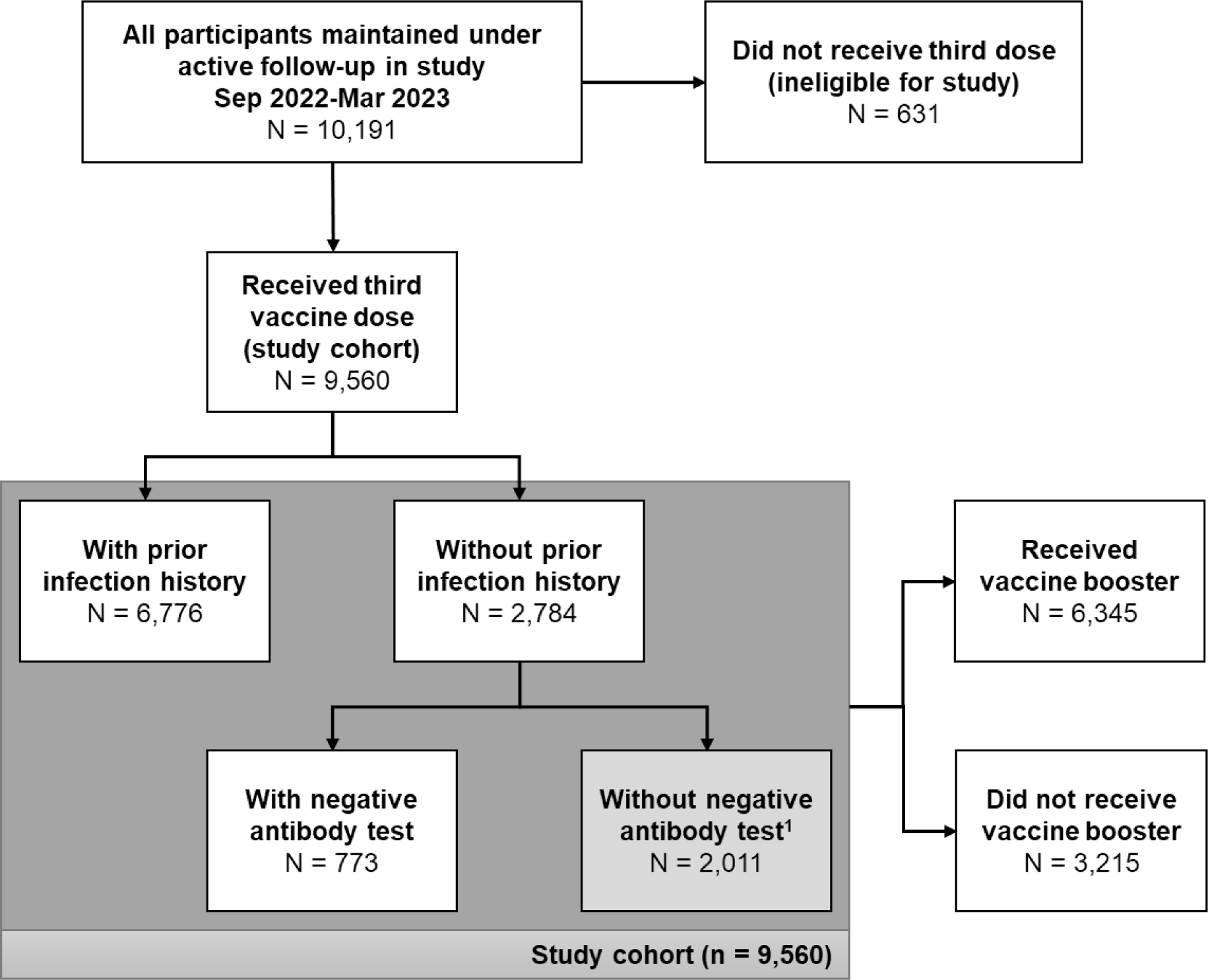
Flowchart of participation in study and uptake of booster vaccine. ^1^Individuals without prior infection history and without a negative antibody test within the 6 months prior to study entry were excluded from analyses exploring time since infection.

The study cohort included both clinical (nurses comprised 33% and doctors 12% of the cohort) and non-clinical, predominantly office-based, roles (administrative/executive staff comprised 17% of the cohort), with 84% being patient-facing overall. The majority (84%) were female, with 88% between the ages 35 to 64, and 90% of white ethnicity. Most (74%) had no chronic medical conditions, although 2.6% reported immunosuppression (Table S3).

Healthcare workers were recruited from every UK region, for this analysis the greatest proportion were resident in Scotland (1,701 participants, 18%), with London (1,199 participants, 13%) and the East of England (1,176 participants, 12%) contributing the greatest proportions from England. Compared to the UK population, participants lived in less socio-economically deprived areas on average (29% in least deprived quintile, 9% in most deprived quintile).

### Crude PCR positivity rates

Over the study period, 1,264 (13.2%) participants received at least one positive PCR result. Including re-infections, we observed 1,298 distinct PCR positives over 1,521,928 person-days of follow-up, corresponding to a crude (unadjusted) PCR positivity rate of 8.53 (95% CI 8.07 to 9.01) per 10,000 days follow-up. Crude PCR positivity rates varied over the analysis period, and by region, but did not differ substantially according to other demographic characteristics (Table S4, Figure S5). Where sequencing information was available, >75% of infections were Omicron BQ.1, BA.5, and XBB (Figure S6).

### Vaccine effectiveness

VE of the fourth dose relative to protection at least six-months after a third dose was estimated as 13.08% (95% CI 0.89 to 23.76) over the entire analysis period. VE was highest in the two months post-vaccination at 23.97% (95% CI 8.48 to 36.83), reducing to 10.30% (95% CI -11.40 to 27.78) in the period two-to-four months post-vaccination, and 1.71% (95% CI -16.97 to 17.40) in the period four-to-six months post-vaccination (Table 1, Figure 2).

**Figure 2:**
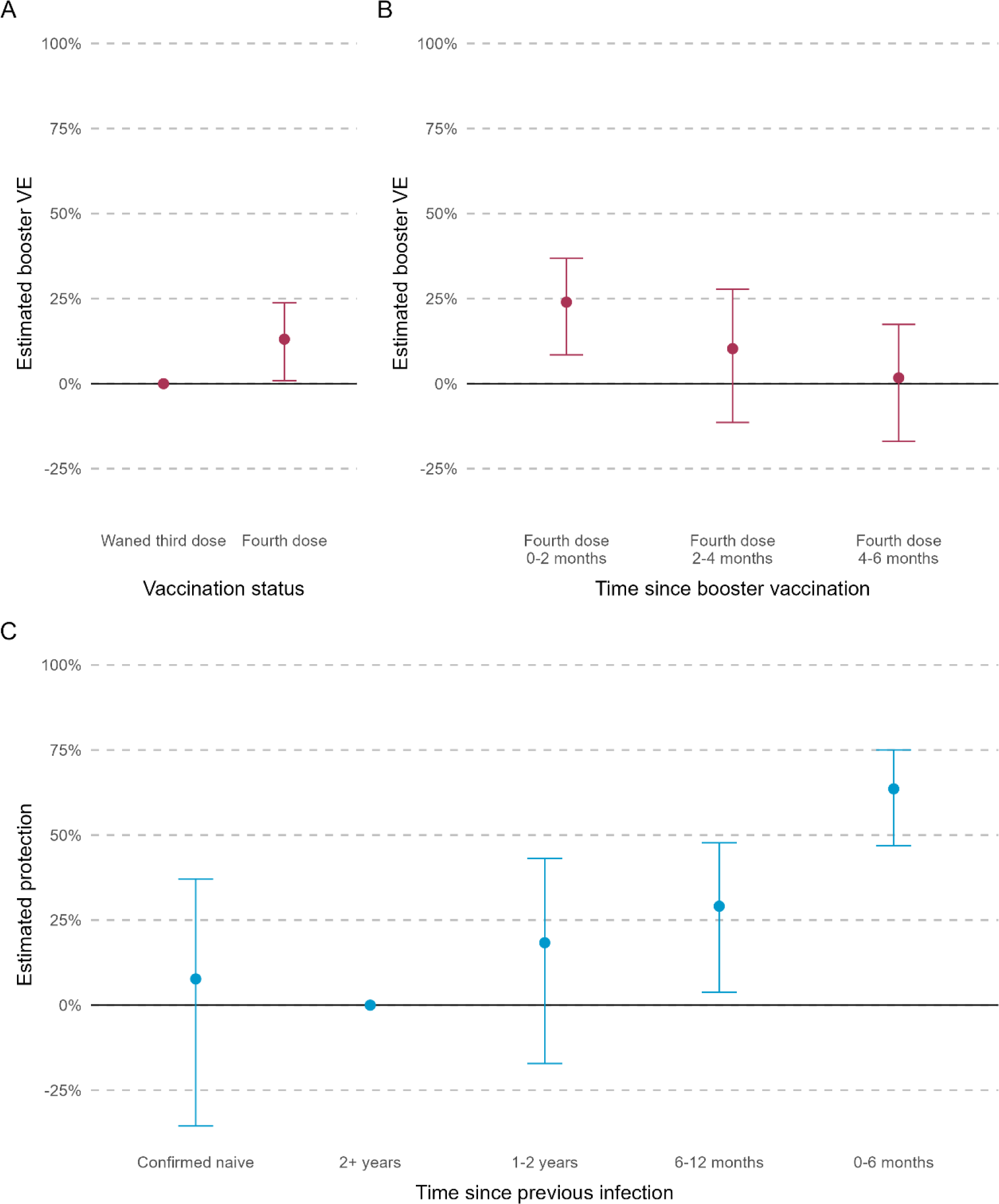
Estimated booster vaccine effectiveness (VE), relative to waned third dose, by booster vaccination status (panel A), time since booster vaccination (panel B), and estimated protection from previous infection, relative to a baseline of 2+ years (panel C). Error bars show the 95% confidence interval around the estimates.

**Table 1:**
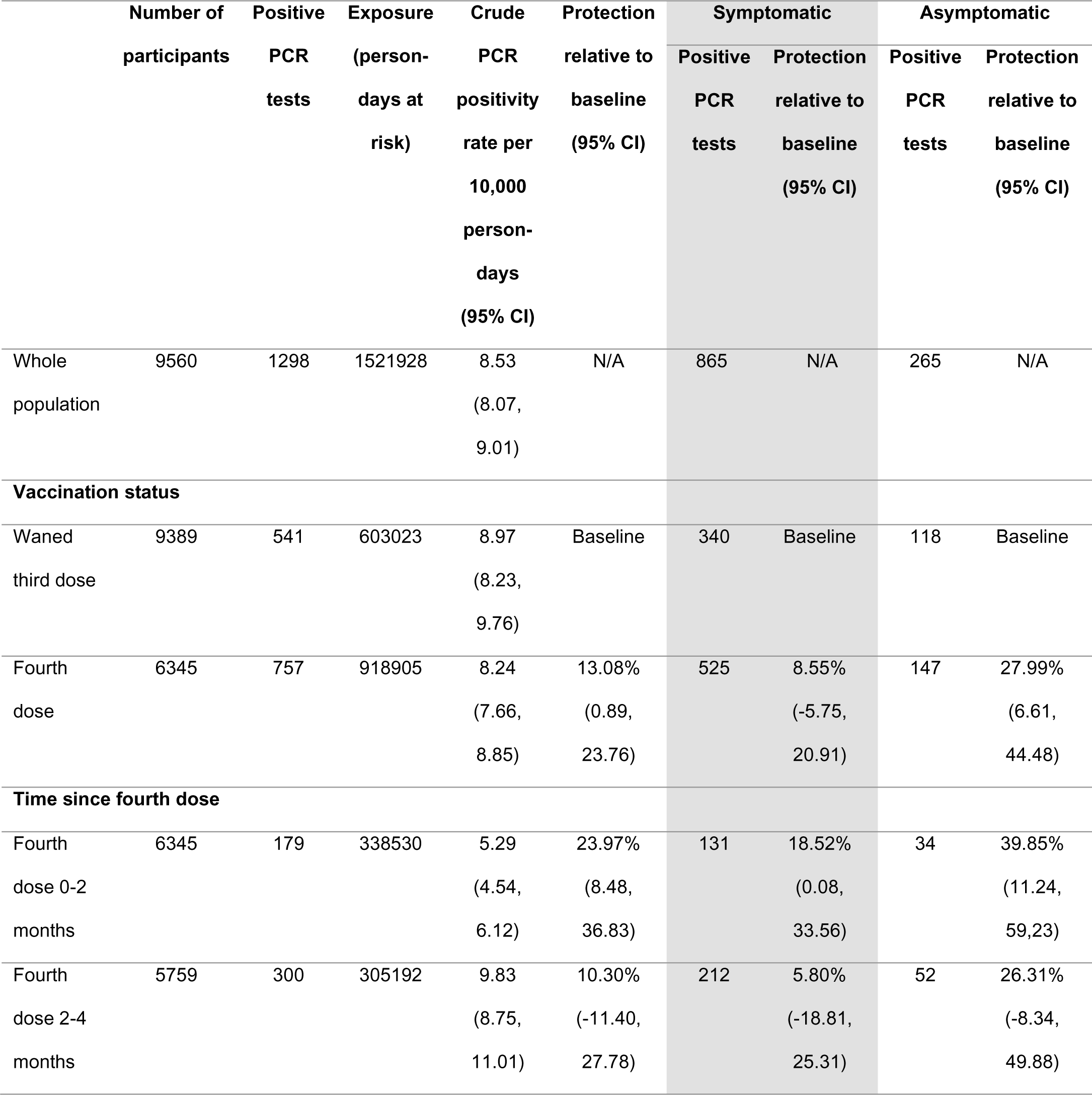
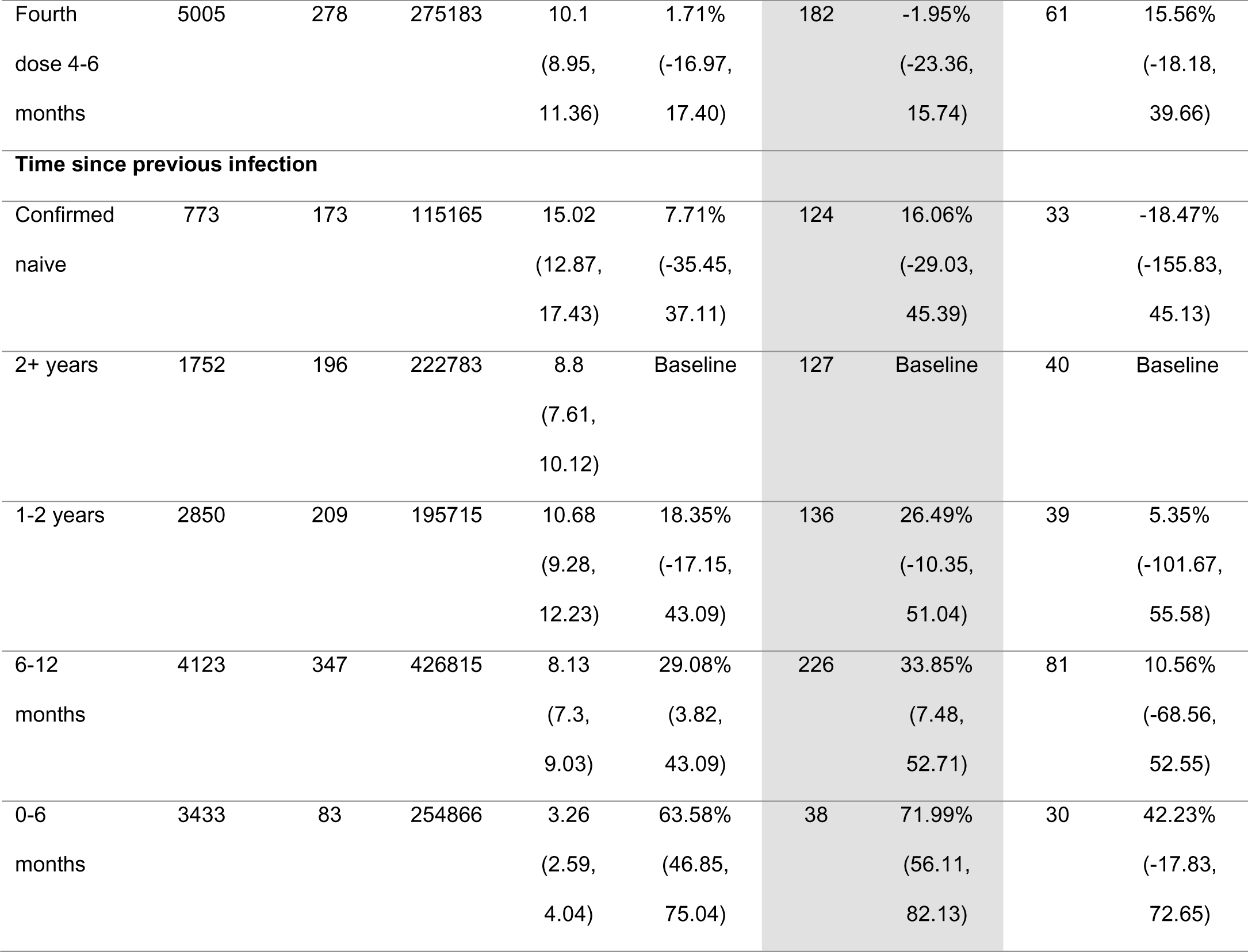
Crude PCR positivity rates per 10,000 person-days and estimated vaccine effectiveness and protection from prior infection by vaccination status, time since previous infection and reported COVID symptoms.

Compared to a waned third dose, VE was 30.87% (95% CI -84.79 to 7.31) among those confirmed as naïve, 32.87% (95% CI 8.51 to 52.20) for those with a previous infection more than 2-years ago, -1.67% (95% CI -40.95 to 26.66) for those with an infection in the past 1-2 years, 23.14% (95% CI 3.07 to 39.06) for infection in the past 6-12 months and 39.58% (95% CI 7.99 to 60.32) for infection in the past 0-6 months (Table S5, Figure S7).

### Protection from previous infection and other covariates

SARS-CoV-2 infection (symptomatic or asymptomatic) between 6 and 12 months ago was associated with a 29.08% (95% CI 3.82 to 43.09) increase in protection compared to individuals who had an infection more than 2 years ago; an infection within the last 6 months was associated with a 63.58% (95% CI 46.85 to 75.04) increase in protection; and those with an infection 1 to 2 years ago or never infected were not associated with significantly more or less protection, 18.35% (95% CI -17.15 to 43.09) and 7.71% (95% CI -35.45 to 37.11), respectively (Table 1, Figure 2).

### Symptomatic vs asymptomatic infection

Among the 1,298 positive PCR results, 1,130 had symptom information reported. Of these, 865 (76.5%) had COVID-19 symptoms, and 265 (23.5%) were asymptomatic (132 had symptoms unrelated to COVID-19, and 133 had no symptoms). The proportion symptomatic was slightly higher among those with a booster dose (78.1%, 525/672) compared to those with a waned third dose (74.2%, 340/458). Among those with an infection in the past 6 months, 55.9% (38/68) reported symptoms compared to >70% of those confirmed naïve or with an infection 6+ months prior (Table 1, Figure S8).

Relative to a waned third dose, VE was 8.55% (95% CI -5.75 to 20.91) against symptomatic infection and 27.99% (95% CI 6.61 to 44.48) against asymptomatic infection. Relative to an infection 2+ years previously, an infection in the past 0-6 months was associated with 71.99% (95% CI 56.11 to 82.13) increased protection against symptomatic infection and 42.23% (95% CI -17.83 to 72.65) against asymptomatic infection (Table 1, Figure 3).

**Figure 3:**
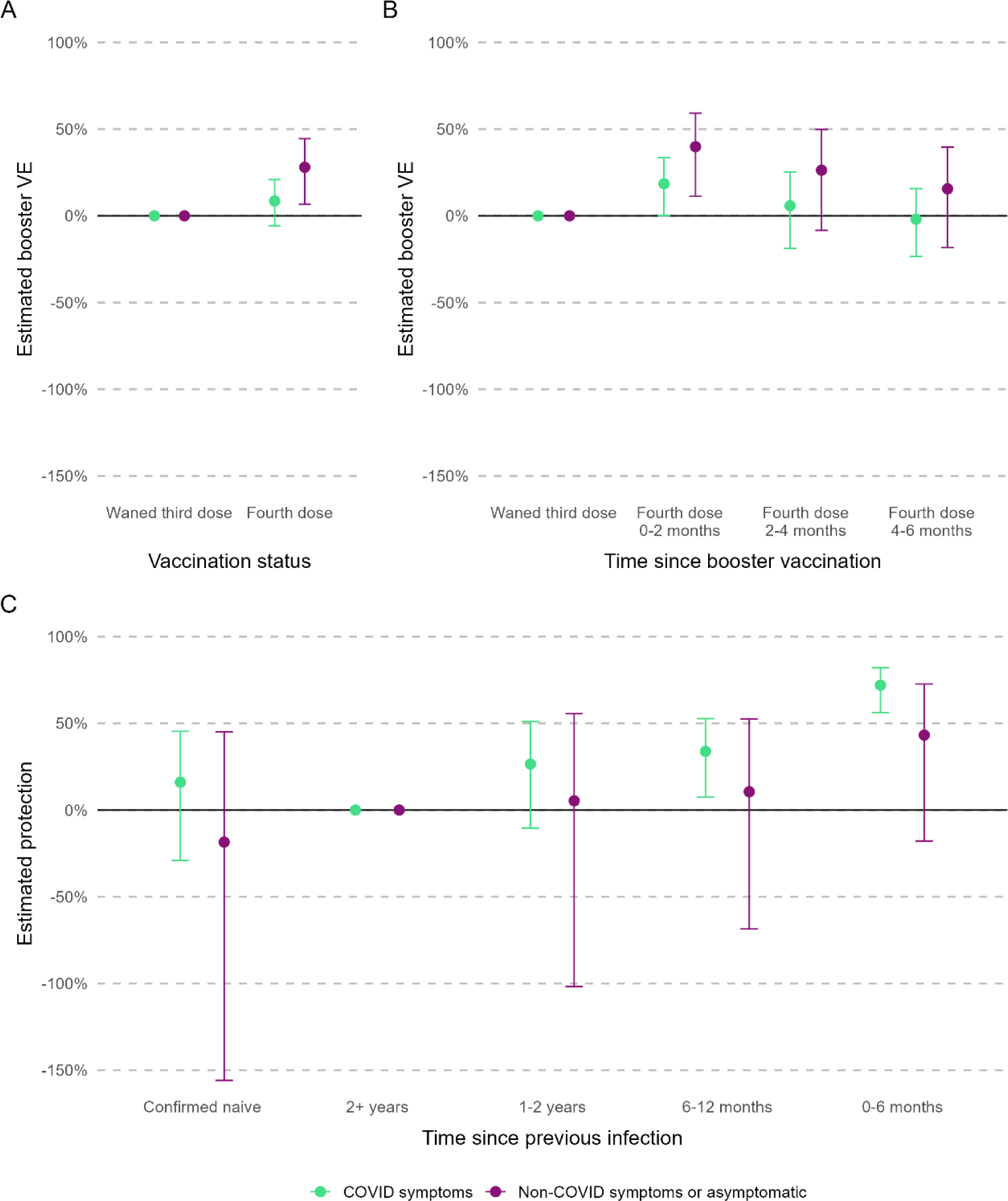
Estimated booster vaccine effectiveness (VE), relative to waned third dose by symptom status and booster vaccination status (panel A), time since booster vaccination (panel B), and estimated protection from previous infection, relative to a baseline of 2+ years (panel C). Error bars show the 95% confidence interval around the estimates.

### Duration of PCR positivity

Duration of PCR positivity was estimated as 7.51 days (95% CI 6.94 to 8.13) overall. When averaged over the study population, this duration was shorter among those with a booster vaccination at 6.90 days (95% CI 5.87 to 8.11), compared to 8.50 days (95% CI 6.79 to 10.64) for those with a waned third dose (Table S6).

The estimated PCR positive duration was 9.51 days (95% CI 7.08 to 12.78) for confirmed naïve participants and 7.25 days (95% CI 6.32 to 8.31) for those with an infection within the past 0-6 months. Infections reported as symptomatic had a longer duration at 8.09 days (95% CI 7.36 to 8.90), compared to 4.70 days (95% CI 4.04 to 5.47) for asymptomatic (Figure 4).

**Figure 4:**
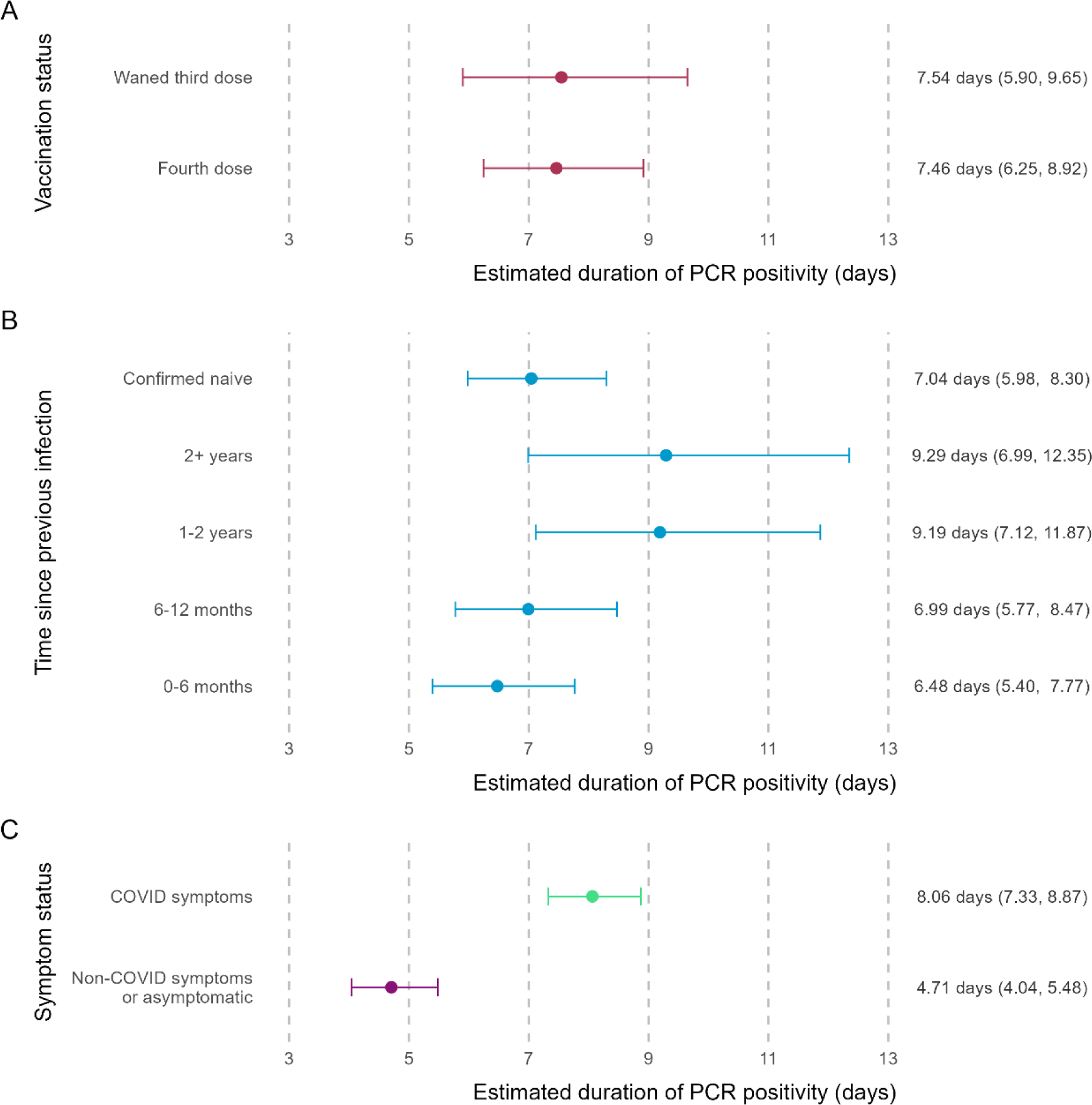
Estimated duration of PCR positivity (days), averaged across the study population, by vaccination status (panel A), time since previous infection (panel B), and symptom status (panel C). Error bars show the 95% confidence interval around the estimated duration of PCR positivity. Estimated time and 95% confidence interval are shown alongside.

## Discussion

We have estimated real-world effectiveness of second COVID-19 booster vaccines, protection against symptomatic and asymptomatic SARS-CoV-2 following previous infection, and the duration of PCR positivity in our cohort of UK healthcare workers over autumn/winter 2022-23, a period with high circulation of Omicron sub-variants. In this cohort of triple-vaccinated, generally healthy, working-age adults we found booster vaccines provided modest and short-lived additional protection against infection. A recent previous infection provided more sustained protection but waning over time was still evident. Asymptomatic infection was more common amongst those with a recent previous infection, and these infections had shorter duration of positivity.

Our study adds to the growing literature on booster VE in the context of high population immunity and high infection rates. A recent cohort study in the Netherlands estimated fourth booster VE of 14% (95% CI 1 to 25%), albeit using self-reported infection data for an older cohort [17]. Meanwhile, early fourth booster VE estimates from a study of United States (US) pharmacies were 49% (95% CI 41 to 55%) and 40% (95% CI 28 to 50%) for age groups 18-49 and 50-64, respectively [18]. This analysis did not control for prior infection history, and biases in the study design mean this may be an over-estimate [19]. In both studies, only data on symptomatic infection was able to be collected, whereas a key strength of SIREN is the capability to detect both symptomatic and asymptomatic infection, and to link to an individuals’ complete testing history.

As seen following third dose vaccination in our cohort [7] and elsewhere [20], protection from infection and protection by vaccination and infection (“hybrid immunity”) conferred longer-lasting immunity than vaccination alone. This may stem from the differential cellular immune responses to infection vs. vaccination [21], additionally, mucosal immunity appears to be conferred by infection rather than vaccination [22,23].

For the Omicron BA.4/5 era in particular, greater and more durable protection was reported for infection-experienced individuals as compared to naïve individuals in results from the nationally-representative UK COVID-19 Infection Survey (CIS) [24], with rapid waning of mRNA vaccine protection from two months post-vaccination. The CIS study estimated >80% protection against Omicron BA.4/5 reinfection for those with a previous Omicron BA.2 infection [24]. This is higher than our estimated 64% protection from a recent (Omicron-period) infection and may reflect our younger cohort and greater number of prior infections among healthcare workers.

A quarter of infections detected in our study were reported as asymptomatic, similar to other European countries during Omicron BA.1 dominance [25]. We found a fourth dose was associated with greater protection against asymptomatic than symptomatic infection, whilst prior infection provided more protection against symptomatic infection, particularly if an infection had occurred recently (i.e. during the Omicron-circulating period). Given most comparably scaled studies have used symptomatic infection as their outcome we are unable to directly compare this result.

We estimate important distinctions in duration of PCR positivity between sub-groups. Several studies have investigated duration of positivity for Omicron-era infections [26] and, whilst most estimate ∼7 days, consistent with our findings, these studies may suffer from small sample sizes or employ methodology that over-estimates duration. In comparison, our analysis correctly accounts for interval-censoring and uncovers important distinctions between sub-groups that would be missed by an empirical approach (e.g. median time between initial PCR positive to subsequent PCR negative). We did not collect information on infectiousness (evidence for which remains varied [26]) but it was notable that individuals with asymptomatic infection, and, to a more limited extent, those either vaccinated or with recent previous infection, were estimated to have shorter durations of PCR-positivity. These estimates can help to inform infection-control measures and transmission models to forecast prevalence.

### Limitations

The SIREN cohort is a cohort of working-age healthcare staff, with participants being predominantly female, of white ethnicity, healthy, and middle-aged. We have controlled for many of these factors in our analysis of VE, however the relatively small proportions of males, older participants, and those with high multimorbidity limits full generalisability to the wider UK population.

Vaccination was not randomly assigned, and despite limited differences in vaccine uptake by measured demographic, we could not control for several other prognostic factors which may be associated with vaccination, e.g. an individual’s perceived exposure risk, which may alter their decision to receive a booster vaccination.

We did not investigate severe disease, which is rare in this cohort. Other studies have found VE against severe disease in the Omicron era to be higher and longer-lasting than against mild disease [27,28].

Given the very small number of unvaccinated individuals in SIREN and recognising they may have different risk profiles to vaccinated individuals, we were unable to use them as a reference group to estimate absolute VE. Previous studies of this cohort have demonstrated a dramatic reduction in infection risk for vaccinated as compared to unvaccinated individuals [29].

We did not compare vaccination type as most of our cohort received the same schedule before study entry (93% three mRNA doses). The COV-BOOST trial found more durable immune response with heterologous third doses [30]. Therefore, potentially the 6% of participants with two viral vector vaccine doses followed by an mRNA vaccine may have had slightly more durable protection, although the effect is expected to be minimal across such a large cohort.

Due to the small number of truly asymptomatic cases, for our symptoms analysis we grouped together asymptomatic cases and those reporting only non-COVID-19-specific symptoms, such as fatigue or muscle ache. Whilst non-specific symptoms occurring around the time of COVID-19 infection may be linked to the infection, this grouping reflects the fact that most participants reported one or more of these non-specific symptoms at some point during the study period, regardless of PCR status.

### Conclusions

In this highly vaccinated, infection-experienced, working-age cohort there was a small but short-lived increase in protection against SARS-CoV-2 infection associated with receipt of booster vaccination during an Omicron sub-variant period. Given the more marginal benefit in comparison with first boosters, and the notably lower coverage of second boosters, economic evaluation will be increasingly important in informing future vaccine deployment. With SARS-CoV-2 yet to settle into a seasonal pattern and considering the short-lived protection provided by current COVID-19 vaccines, it appears premature to plan mass roll-out of annual boosters akin to Influenza. Currently, therefore, vaccine boosters remain an important tool in responding to the dynamic COVID-19 landscape; boosting population immunity in advance of periods of anticipated pressure, such as surging infection rates or emerging variants of concern.

## Supporting information

SIREN Study Group

## Acknowledgements

We thank all the participants for their ongoing contributions and commitment to this study; the research teams at all 89 SIREN sites actively following up participants during this analysis period for their support and for making the study possible; colleagues at the U.K. Health Security Agency Sero-epidemiology Unit, namely Ezra Linley and Simon Tonge, for their support in biobanking and processing the high volumes of serum samples; and colleagues at the U.K. Health Security Agency Porton Down for organizing and performing all the centralized serologic testing, including testing of 35,000 baseline samples. We thank Shaun Seaman and Brian Tom at the MRC Biostatistics Unit, University of Cambridge for helpful discussion.

## Ethics approval

The study protocol was approved by the Berkshire Research Ethics Committee on May 22, 2020. This study collected consent from all participants. ISRCTN Registry number: ISRCTN11041050.

## Participant and public involvement

The SIREN study has an active Participant Involvement Panel (PIP), co-delivered with the British Society for Immunology. The SIREN PIP meets regularly to consult participants on proposed study developments, recent outputs and obtain feedback on study delivery. We have a multimethod cohort retention programme informed by the PIP and wider participant feedback which we collect continuously and through cross-sectional surveys.

## Dissemination to participants and related patient and public communities

We use diverse channels to disseminate our findings to participants, including via regular digital newsletters, live webinars, videos, blogs and through our study webpage on the UKHSA website (https://www.gov.uk/guidance/siren-study).

UKHSA and the MRC Biostatistics Unit have public facing websites and Twitter accounts @UKHSA and @MRC_BSU. UKHSA and the MRC Biostatistics Unit engage with print and internet press, television, radio, news, and documentary programme makers.

## Transparency statement

The lead author affirms that this manuscript is an honest, accurate, and transparent account of the study being reported; that no important aspects of the study have been omitted; and that any discrepancies from the study as planned (and, if relevant, registered) have been explained.

## Data availability

The metadata will be available through the Health Data Research UK Co-Connect platform and will be available for secondary analysis once the study has completed reporting.

## Code availability

A list of all R packages used and R code written to process the data, implement the statistical analysis, and produce the figures and tables is available online at https://github.com/SIREN-study/SARS-CoV-2-booster-vaccination.

## Contributors

PDJ, VJH, SF, and SH designed the analysis plan and wrote the paper. VJH and SF cleaned and finalised the dataset for analysis and did the descriptive analyses. PDK, AC, AP, and DDA planned and did the statistical analysis. VJH, SF, AC, and AA verified the data in the study. All authors reviewed and approved the manuscript for publication. All authors had full access to all the data in the study and accept responsibility to submit for publication.

