## Supplementary Materials for "Protection of second booster vaccinations and prior infection against SARS-CoV-2 in the UK SIREN healthcare worker cohort"

### Supplementary Appendix for: Protection of COVID-19 booster vaccination and previous infection against symptomatic and asymptomatic SARS-CoV-2

**2 Supplementary figures** **8**

**3 Supplementary tables** **17**

**References** **25**

### 1 Supplementary Methods

#### 1.1 Multi-state models

##### Overview

Multi-state models can be used to describe health-related processes over time, particularly those relating to disease epidemics. In multi-state models, an individual begins in a given state, spends a period of time in that state, then moves to a next state. Examples include survival models, where individuals move from the alive state to the dead state, and disease-staged models, where each state represents disease severity, with movement from state to state representing disease progression [1]. The period of time spent in a state and the state to which the individual moves can be thought of as random variables which may be estimated.

##### Generic multi-state model

Multi-state models are governed by a set of transition intensities $q_{rs}(t, z(t))$ for each pair of states $r$ and $s$. The intensities may also depend on the time of the process $t$, or more generally a set of individual-specific and/or time-varying explanatory variables $z(t)$.

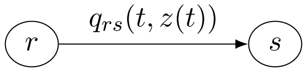

The transition intensity represents the instantaneous risk of moving from state $r$ to state $s$ and is defined by the limit as $\delta t$ tends to zero:

$$q_{rs}(t, z(t)) = \lim_{\delta t \to 0} \frac{P(S(t + \delta t) = s | S(t) = r)}{\delta t}$$

The intensities form a transition intensity matrix Q whose rows sum to zero, with diagonal entries defined by:

$$q_{rr}(t, z(t)) = -\sum_{s \neq r} q_{rs}(t, z(t))$$

Typically multi-state models will be Markov, i.e. with the assumption is that future evolution only depends on the current state i.e. $q_{rs}(t, z(t), \mathcal{F}_t) = q_{rs}(t, z(t))$, where $\mathcal{F}_t$ is the observation history of the process up to time $t$. For conciseness, hereon we denote the implicitly time-varying transition intensity simply as $q_{rs}$.

Let $Q$ be the $r \times s$ matrix formed by the $q_{rs}$, then the transition probability matrix, with probability of moving between states in time $t$, $P(t)$ is:

$$P(t) = \exp(tQ) = \sum_{n=0}^{\infty} \frac{t^n}{n!} Q^n$$

and the sojourn time (mean time spent in a state) is:

$$E(T_r) = -\frac{1}{q_{rr}}$$

##### Covariates

Covariates can be included in multi-state models through a proportional hazards model, whereby the transition intensities $q_{rs}$ are replaced by $q_{rs}(\boldsymbol{z_i})$ for a given set of $m$ constant or time-varying covariates $z_{im}$:

$$q_{rs}(z_i) = q_{rs}^{(0)} \exp\left(\sum_{m=1}^{M} \beta_m z_{im}\right)$$

where $q_{rs}^{(0)}$ are the baseline hazards and $\exp(\beta_m)$ is the hazard ratio for the $m$th covariate.

##### Censoring

Multi-state models are well-suited to investigating interval-censored 'panel' data, where observations represent PCR testing rather than the occurrence of infection (i.e. where sampling times are non-informative). The time intervals of observation for the SIREN study are fixed in advance (2-weekly intervals). A continuous-time model is appropriate in the context of these data, allowing for changes of state (from susceptible to infected, and infected to recovered) to occur at any point during the 2 week interval. This multi-state framework additionally allows incomplete testing (i.e. gaps in an individual's testing history) to be implicitly accounted for, without the assumption that an individual remains uninfected despite not testing.

For an individual with a PCR negative status at time L and PCR positive status at time R, infection is assumed to occur at some time $T \in [0, \infty)$ during the time interval $(L, R]$. The infection time is interval-censored, $T \in (L, R]$, and, if the time intervals are fixed in advance, this interval censoring is independent of $T$ [2]:

$$P(L < T \leq R | L = l, R = r) = P(l < T < r)$$

##### Likelihood

For interval-censored panel data we have information on states $(x_{i1}, \ldots, x_{in_i})$ at times $(t_{i1}, \ldots, t_{in_i})$ for person $i$ which may take the values PCR negative (Susceptible) or PCR positive (Infected). As described by Kalbfleisch & Lawless [3], the likelihood contribution to the transition intensities $q_{rs}$ from person $i$ is:

$$L_i(q_{rs} \mid x_i) = p(x_{i1} \mid x_{i0}) p(x_{i2} \mid x_{i1}) \ldots p(x_{in_i} \mid x_{in_i-1})$$
$$\text{where: } p(x_{ij} \mid x_{ij-1}) = p_{x_{i,j-1}, x_{i,j}}(t_{i,j-1}, t_{i,j} \mid q_{rs})$$

The full likelihood $L(Q)$ is the product of the $L_{i,j}$ over all individuals and transitions. An estimate of the transition intensity matrix $Q$ is obtained by maximising this likelihood.

##### Piecewise-constant hazards

Risk of community transmission of COVID-19, as well as risk of within-hospital transmission for healthcare workers, has varied throughout the study. The calendar time at which a SARS-CoV-2 infection takes place must therefore be considered. A time-varying covariate $z(t)$ for calendar month allowing for piecewise-constant hazards for each month are used to model the changing risk of infection over the study period.

##### Model specification and implementation

Two multi-state models were used for this study:

The first was a two-state model where participants were grouped into susceptible (with a negative PCR test) or infected (with a positive PCR test).

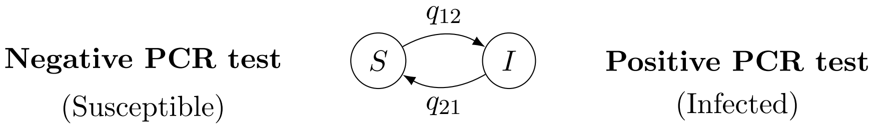

The second model included symptom status information as an outcome, sub-dividing the infected state into asymptomatic or symptomatic infection.

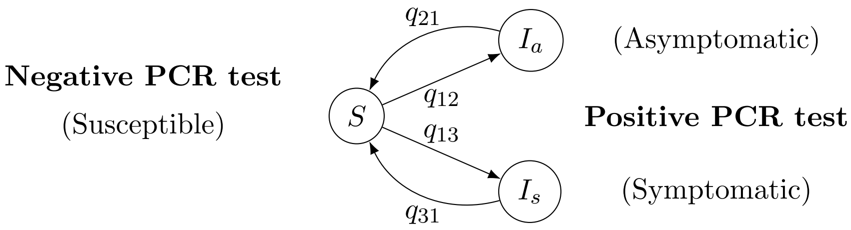

The models were specified to include proportional hazards for selected covariates on each transition and piecewise-constant hazards for the month covariate to capture the changing risk of SARS-CoV-2 infection (Table S1).

The R package msm [4] was used to implement maximum-likelihood estimation for these multi-state models.

#### 1.2 Cox proportional hazards models

##### Overview

The Cox proportional hazards models is another common approach used to fit regression models with survival outcomes.

Following the notation used previously we define a hazard (which may be time-dependent) at time $t$ as $q(t)$, with a baseline hazard $q^{(0)}(t)$, then for a given set of $m$ constant or time-varying covariates $z_{im}$:

$$q(t|z_i) = q^{(0)}(t) \exp\left(\sum_{m=1}^{M} \beta_m z_{im}\right)$$

where $\exp(\beta_m)$ is the hazard ratio for the $m$th covariate.

##### Stratified Cox proportional hazards model

A key assumption of Cox models is that hazards are proportional. For covariates which exhibit non-proportionality, covariate stratification may be used.

For the example of vaccination and age, an unstratified model would state:

$$q(t|z) = q^{(0)}(t)\exp(\beta_{\text{age}} z_{\text{age}} + \beta_{\text{vax}} z_{\text{vax}})$$

This implies that the trend in survival over time is the same for all age groups, and that (hazard for age group 2)/(hazard for age group 1) is a constant. Stratification avoids needing to make this assumption by allowing the baseline hazard to differ by different covariate levels, fitting two (or more) separate equations with a common regression coefficient $\beta_{\text{vax}}$:

$$q_{\text{age}=1}(z) = q^{(0)}_{\text{age}=1}\exp(\beta_{\text{vax}} z_{\text{vax}})$$
$$q_{\text{age}=2}(z) = q^{(0)}_{\text{age}=2}\exp(\beta_{\text{vax}} z_{\text{vax}})$$

##### Comparison of estimated hazards

Two MSMs were used for the main analysis: the two-state model where participants moved between 'susceptible' (PCR-negative) and 'infected' (PCR-positive) states, and the three-state model which incorporated symptom status in the 'infected' states. For comparison purposes, a stratified Cox proportional hazards model was fitted to the same dataset. The estimates of this simpler model (which didn't consider recovery or interval censoring) were able to be compared to the more complex models, as well as providing a historical comparison to previous SIREN analyses which have used stratified Cox models only.

We used three models to generate our estimates: the Cox model (equivalent to a two-state MSM without recovery or interval censoring), the two-state MSM with recovery, and the three-state MSM with recovery. Each of the three models was run with four scenarios:

1. Not including time since previous infection as a covariate and using a binary indicator of vaccination, to estimate overall vaccination effectiveness in a cohort with heterogeneous time since previous infection.

2. Not including time since previous infection as a covariate and using a time-varying indicator of vaccination, to estimate vaccine effectiveness by time since vaccination in a cohort with heterogeneous time since previous infection.

3. Including time since previous infection and a binary indicator of vaccination, to estimate protection associated with prior infection.

4. Including time since previous infection and an interaction effect between this variable and vaccination, to estimate the marginal effectiveness of vaccination by time since prior infection.

The list of models is shown in Table S2, and Figures S1-S4 compare the estimates from the MSM and Cox proportional hazards models.

For the main analysis we converted the hazard ratios (HR) estimated by the MSMs into vaccine effectiveness (VE) and relative protection estimates using the formula: VE = 1 − HR. Duration of positivity estimates were averaged over the study population to provide comparable estimates.

The R package survival [5] was used to implement estimation for the stratified Cox proportional hazards models. The Cox proportional hazards models included the covariates specified previously, with stratification on the occupational setting, age group, and region covariates. As with previous analyses of this cohort, we allowed for deferred study entry and re-entry, with participants entering or re-entering the cohort 6 weeks after the date of a positive PCR test. The Cox models were fitted both with and without a random effect on the hospital trust (i.e. a frailty model) to control for trust-specific effects, with very minimal effect on estimates [6].

##### 1.3 Methodological discussion

###### Use of multi-state models

We estimated very similar hazard ratios from the MSMs and comparable Cox proportional hazards models. The slightly wider confidence intervals from the MSMs may be a more accurate reflection of the uncertainty in the estimates, given that they account for the interval censoring/missed testing.

Using MSMs we could account for gaps due to missed testing and jointly estimate the time spent in a positive state, as well as symptomatic vs asymptomatic infection. Such methods are growing in popularity, with tutorials directed towards epidemiological research [7, 8, 9], and are well-suited to studies involving interval-censored panel-style data, although we note the MSMs took significantly longer to fit in our study (several hours compared to under a minute for the Cox proportional hazards model) and required careful tuning of the optimizer to converge to the maximum likelihood.

Stratification and piecewise-constant hazards over time were used to account for non-proportionality, however, it is possible that the proportional hazards assumption may still be unrealistic for the covariates we have studied, a common issue particularly in medical studies. Stensrud and Hernán recommend that hazard ratios should be interpreted as a weighted average of the true hazard ratios over the entire follow-up period [10].

###### Choice of baseline

For this study we elected to use ‘2+ years’ as the baseline group for the covariate time since previous infection. Whilst a comparison to the immunological baseline (confirmed naïve) would be desirable, in our study this is a very small group and using this baseline introduces greater uncertainty to the estimates. Furthermore, the infection trends for the confirmed naïve group indicate likely differences in risk behaviour, e.g. more shielding, making them less representative of the larger cohort.

Whilst the pandemic has evolved, the SIREN study continues to provide relevant and generalisable results. Most of our study population have now experienced a prior COVID-19 infection, therefore selecting a group with a previous infection as a baseline seems appropriate. We chose ‘2+ years’ due to size of this group, and based on previous analysis [11] it was reasonable to assume this group was the least protected. Our analyses test whether a more recent infection was indicative of better protection.

For completeness, we have re-run the estimates with the confirmed naïve group as a baseline, and include these in Figures S10 and S11.

##### Causal interpretation of VE

Work to investigate VE under different counterfactual scenarios is underway to more fully assess the causal effect of vaccination on reducing acquisition of SARS-CoV-2 infection in this population [12].

#### 2 Supplementary figures

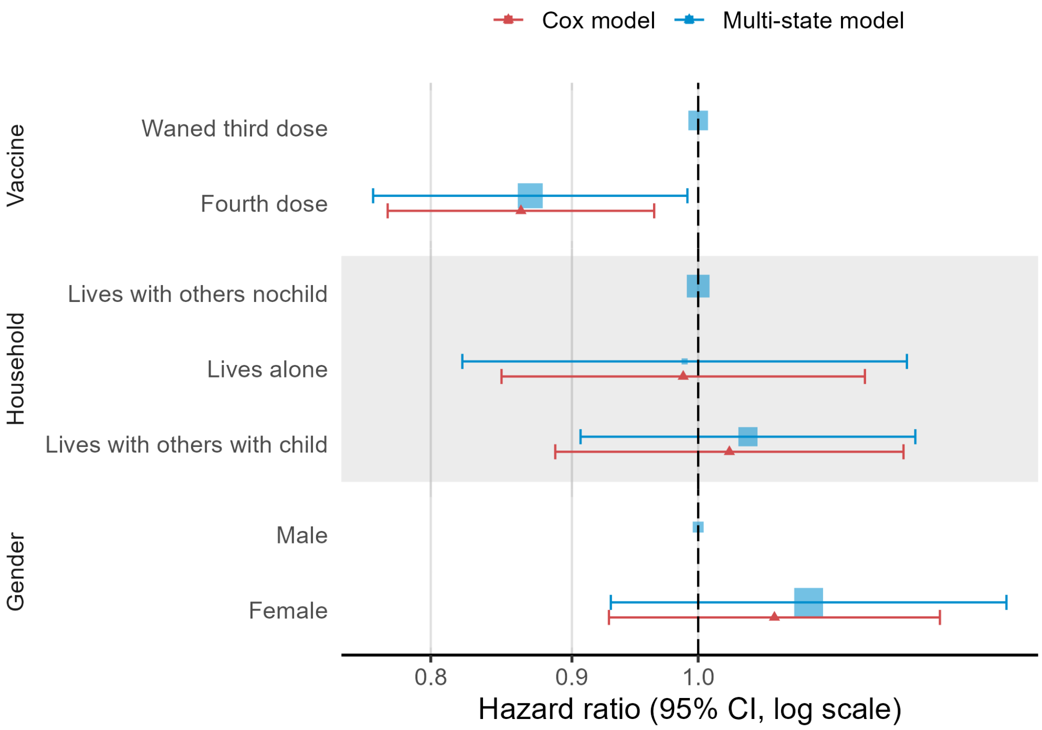

Figure S1: Comparison of hazards estimated by MSM model 1 and Cox model 1, for covariates common between the two models. Error bars show the 95% confidence interval around the estimated hazards, the size of the marker indicates the relative size of each subgroup.

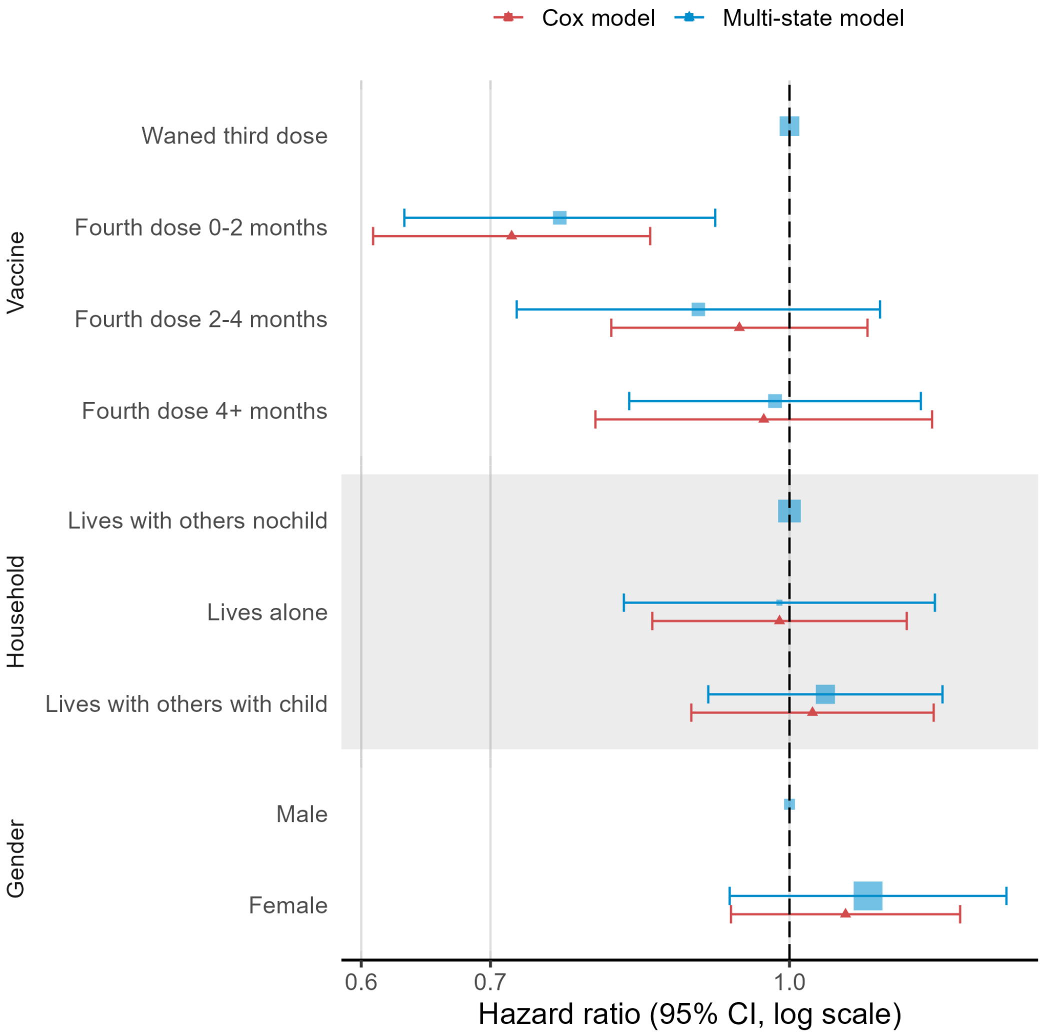

Figure S2: Comparison of hazards estimated by MSM model 2 and Cox model 2, for covariates common between the two models. Error bars show the 95% confidence interval around the estimated hazards, the size of the marker indicates the relative size of each subgroup.

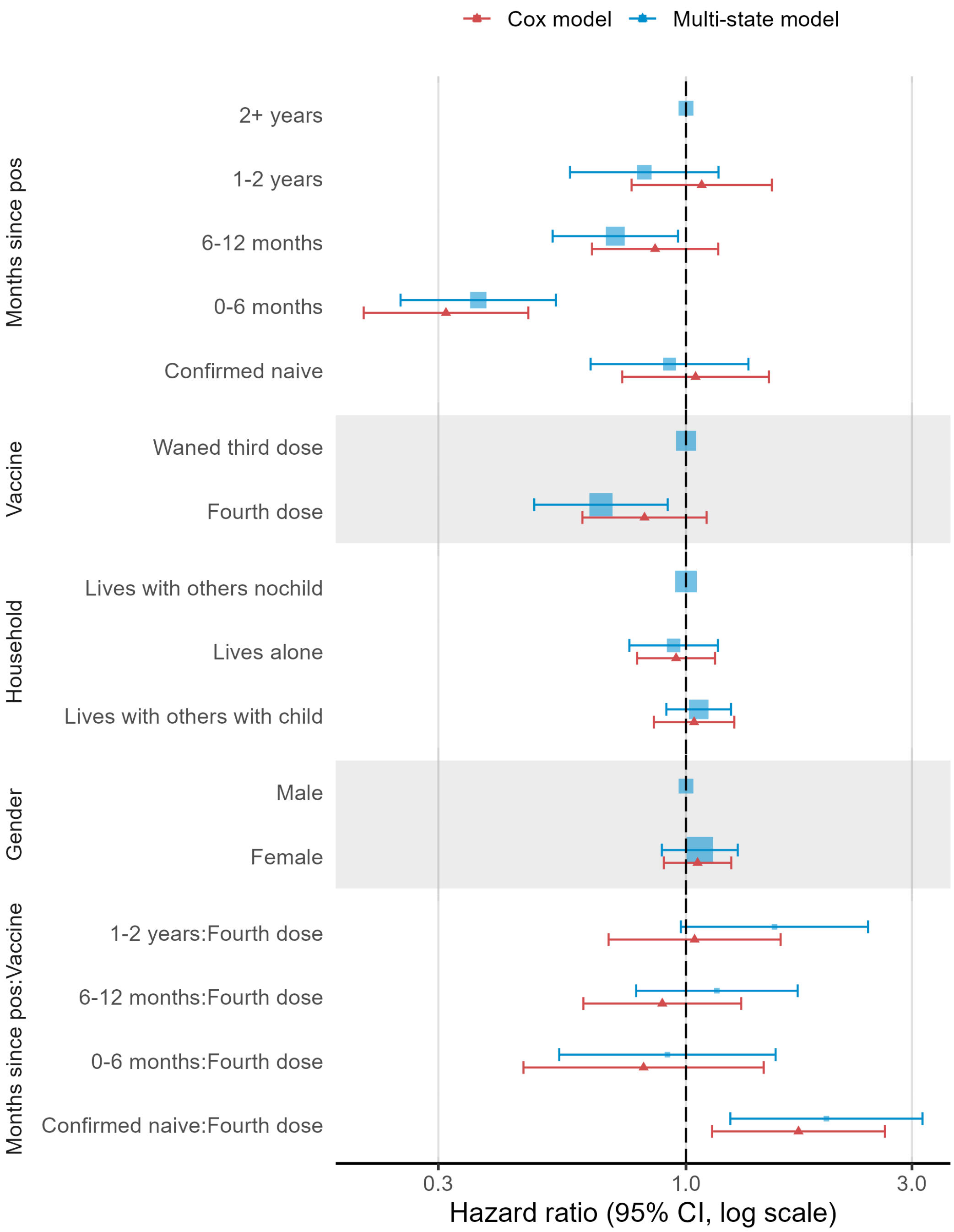

Figure S3: Comparison of hazards estimated by MSM model 3 and Cox model 3, for covariates common between the two models. Error bars show the 95% confidence interval around the estimated hazards, the size of the marker indicates the relative size of each subgroup.

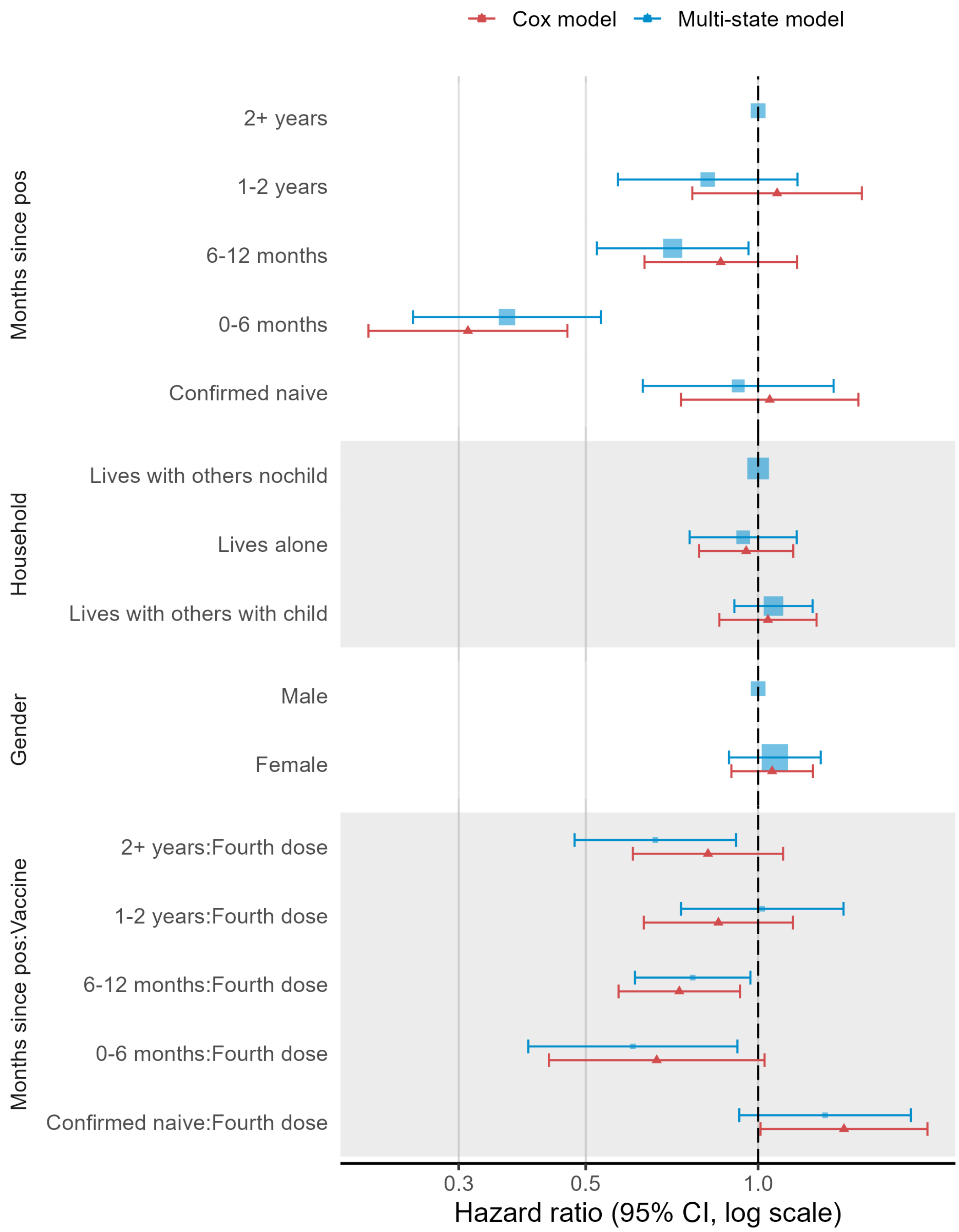

Figure S4: Comparison of hazards estimated by MSM model 4 and Cox model 4, for covariates common between the two models. Error bars show the 95% confidence interval around the estimated hazards, the size of the marker indicates the relative size of each subgroup.

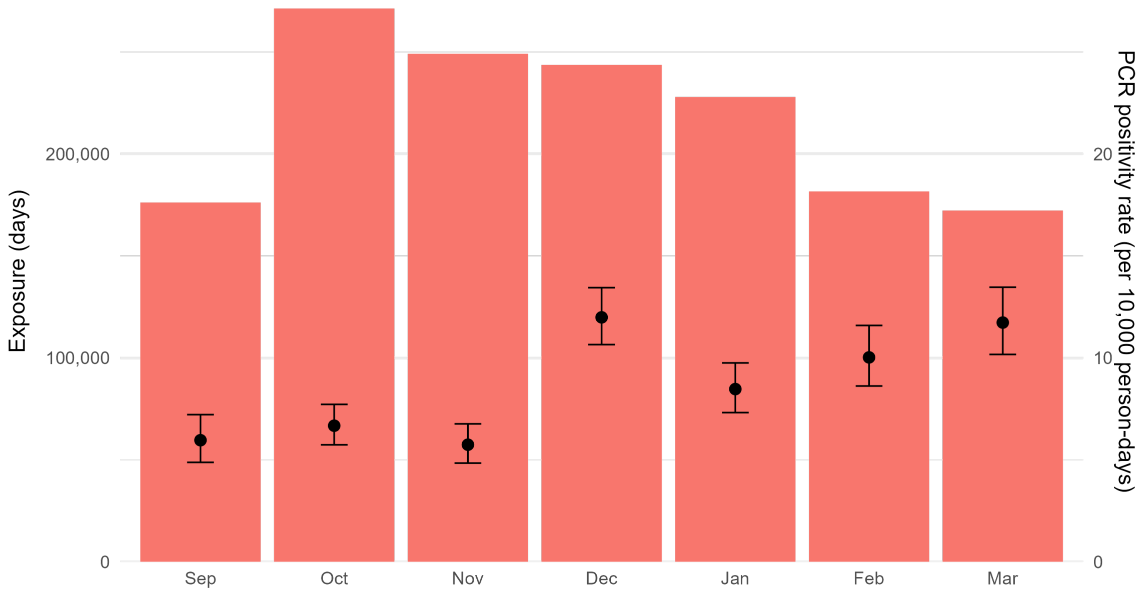

Figure S5: Exposure (person-days at risk) and crude PCR positivity rate per 10,000 person-days by month. Left-hand y-axis for exposure (bar chart), right-hand y-axis for crude PCR positivity (points and error-bars). Error bars show the 95% confidence interval (estimated using an exact Poisson method) around the estimated PCR positivity rate.

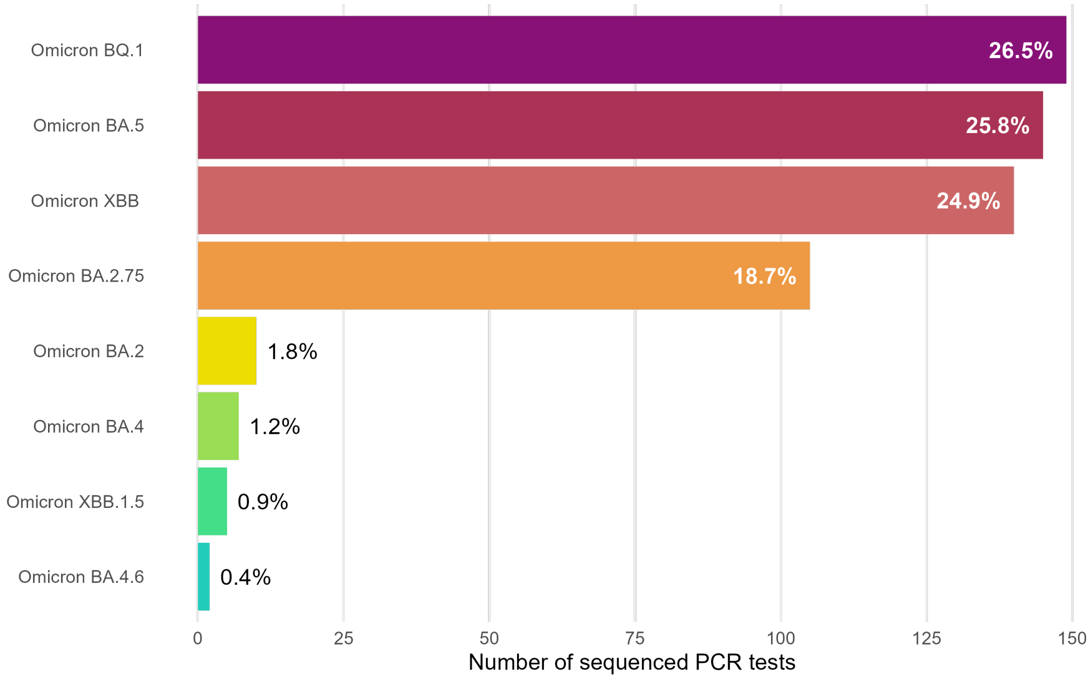

Figure S6: SARS-CoV-2 infections, by sequenced variant. Percentages indicate proportion of sequenced variants. Sequence information was available for 43.4% (563/1,298) of detected infections.

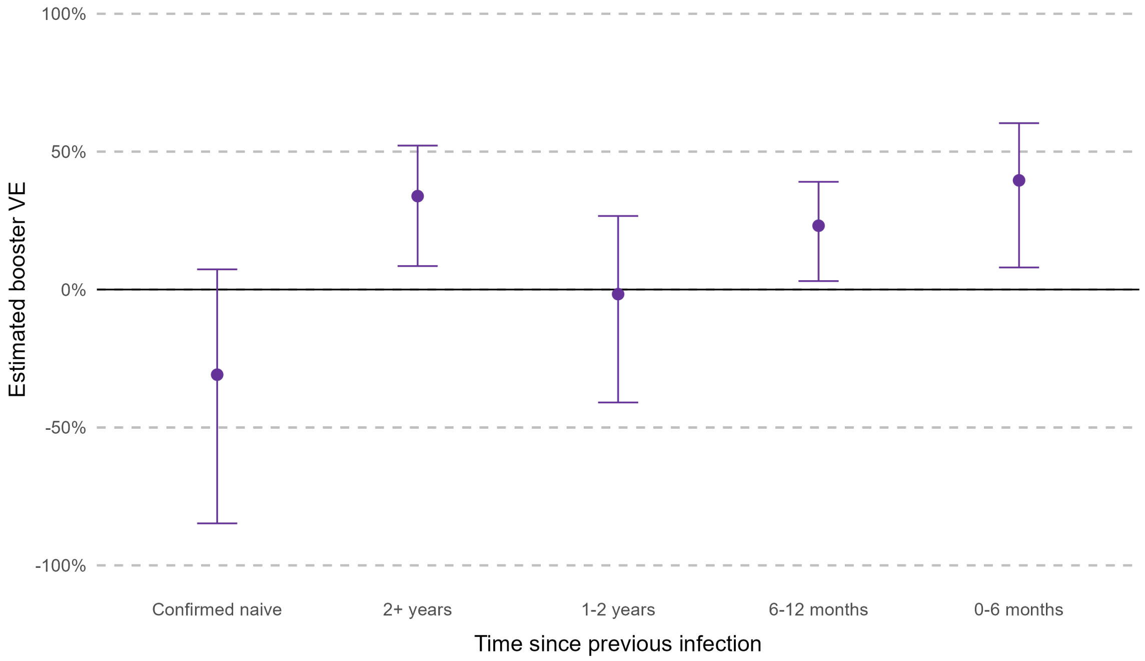

Figure S7: Estimated booster vaccine effectiveness (VE), relative to waned third dose, by time since previous infection. Error bars show the 95% confidence interval around the estimated VE.

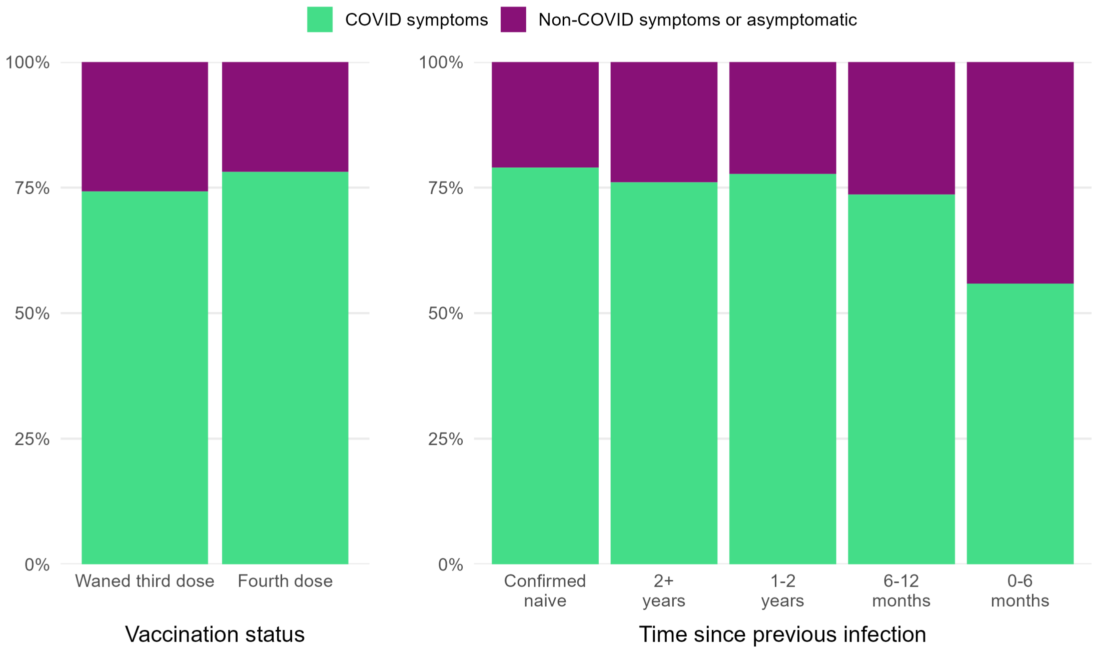

Figure S8: SARS-CoV-2 infections, by vaccination status, time since previous infection, and presence of COVID-19 symptoms.

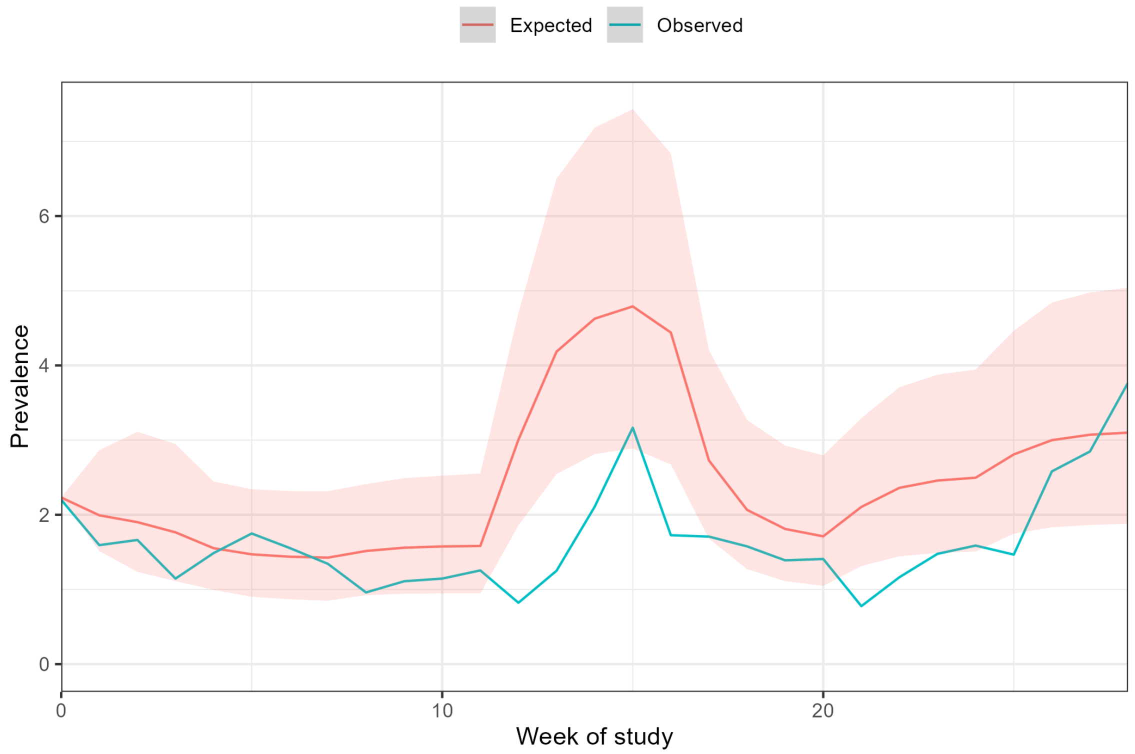

Figure S9: Comparison of expected (estimated) and observed prevalence in the infected state over time. Shaded area shows the 95% CI around the expected prevalence. The observed prevalence is below the lower 95% CI of expected prevalence in weeks 12-16. This period coincides with December and the new year period, when lower adherence to fortnightly testing was observed. This difference may be a result of the model accounting for infections that are unobserved due to the interval-censored nature of the data.

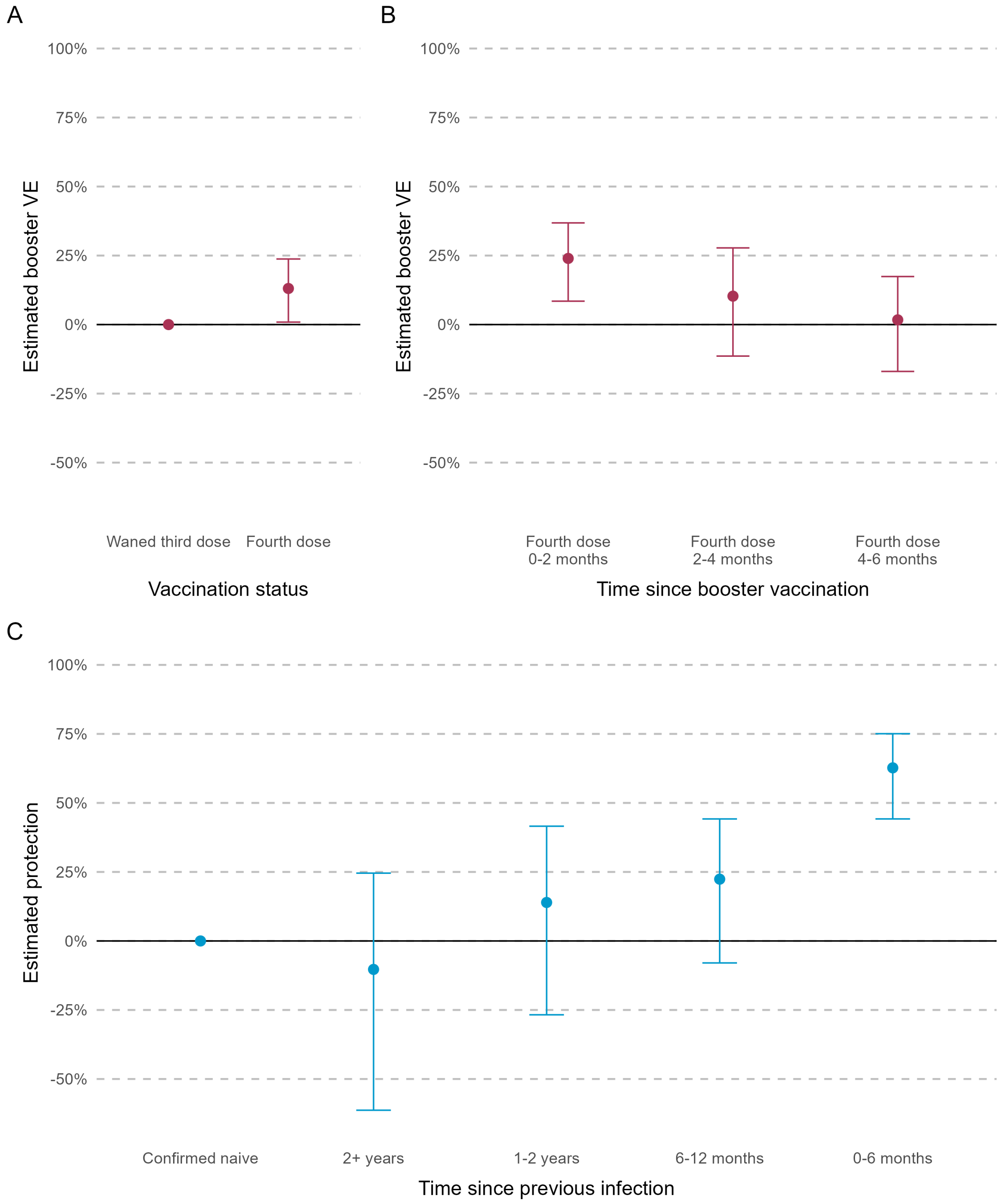

Figure S10: Estimated booster vaccine effectiveness (VE), relative to waned third dose, by booster vaccination status (panel A), time since booster vaccination (panel B), and estimated protection from previous infection, relative to a baseline of confirmed naïve (panel C). Error bars show the 95% confidence interval around the estimates.

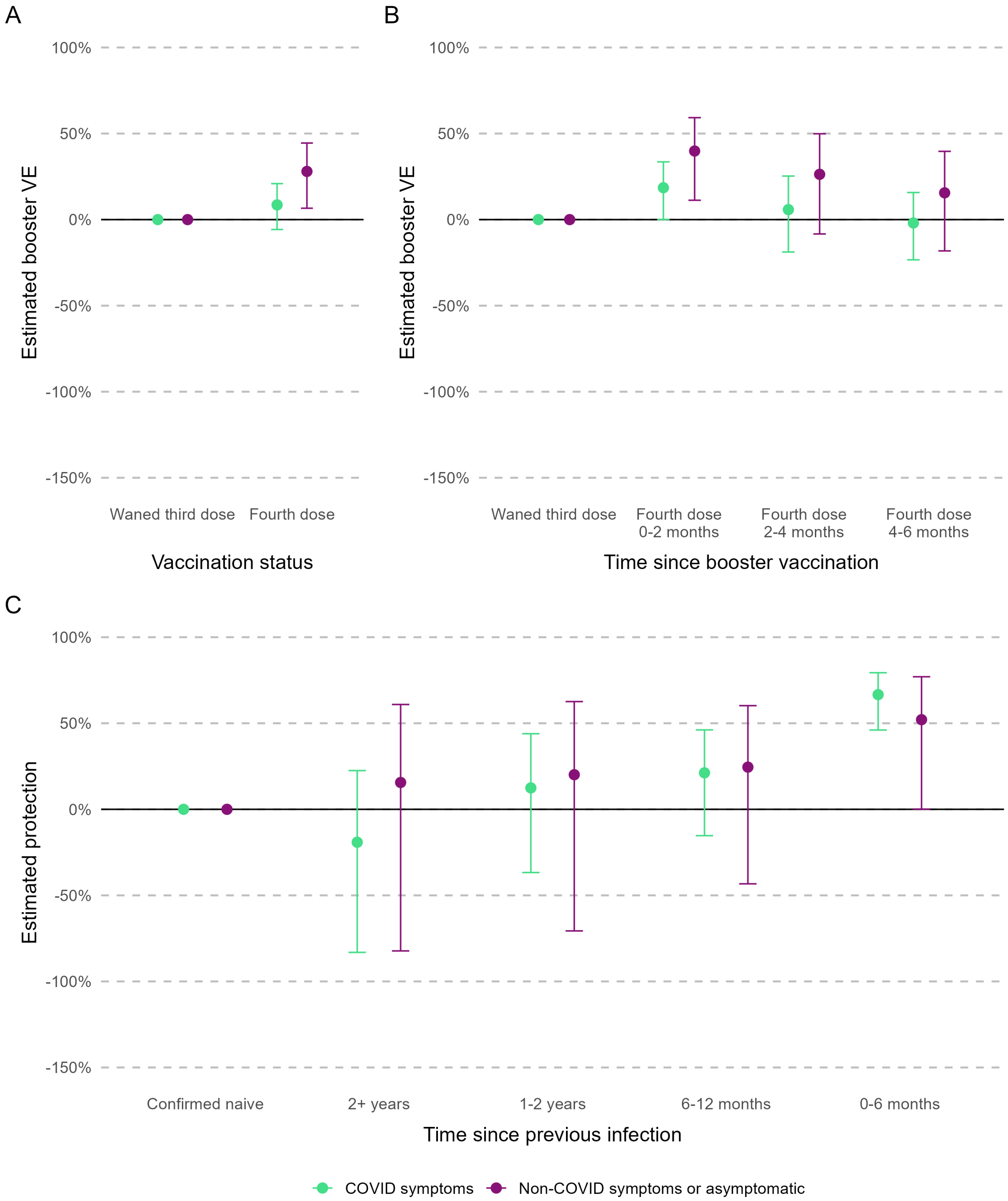

Figure S11: Estimated booster vaccine effectiveness (VE), relative to waned third dose by symptom status and booster vaccination status (panel A), time since booster vaccination (panel B), and estimated protection from previous infection, relative to a baseline of confirmed naïve (panel C). Error bars show the 95% confidence interval around the estimates.

### 3 Supplementary tables

Table S1: List of covariates used for multi-state models, with time-dependence and transitions used for each covariate.

| **Covariate** | **Levels** | **Time-dependent** | **Transitions** |
|---|---|---|---|
| Month | Sep, Oct, Nov, Dec, Jan, Feb, Mar | Yes, piecewise-constant hazards | Infection only |
| Vaccination status | Third dose, Fourth dose (0-2 months, 2-4 months, 4-6 months) | Yes | Infection and recovery |
| Binary vaccination status | Third dose, Fourth dose | Yes | Infection and recovery |
| Time since previous infection | Confirmed naïve, 2+ years, 1-2 years, 6-12 months, 0-6 months | Yes | Infection and recovery |
| Age group | <25, 25-34, 35-44, 45-54, 55-64, 65+ | No | Infection and recovery |
| Gender | Male, Female | No | Infection only |
| Region | East Midlands, East of England, ... | No | Infection only |
| Household status | Lives alone, Lives with others (no children), Lives with children | No | Infection only |
| Occupational setting | Ambulance, ED, Inpatient, Outpatient, ... | No | Infection only |

Table S2: List of multi-state and Cox proportional hazards models used to generate estimates, with number of individuals, inclusion of symptom information, and list of covariates. Individuals without prior infection history and without a negative antibody test were excluded from analyses exploring time since infection, this is indicated by a smaller number of individuals for these models.
x: covariate included as main effect in regression, p: covariate included with piece-wise constant hazards, s: covariate included with stratification.

| **Multi-state models** | MSM model 1 | MSM model 2 | MSM model 3 | MSM model 4 | MSM model 5 | MSM model 6 | MSM model 7 |
|---|---|---|---|---|---|---|---|
| N (individuals in model) | 9,560 | 9,560 | 7,549 | 7,549 | 9,560 | 9,560 | 7,549 |
| Symptom data (Y/N) | N | N | N | N | Y | Y | Y |
| **Covariates** | | | | | | | |
| Month | p | p | p | p | p | p | p |
| Binary vaccination status | x | | x | | x | | x |
| Vaccination status | | x | | | | x | |
| Time since infection | | | x | x | | | x |
| Vaccination:time since infection (interaction term) | | | x | x | | | x |
| Age group | x | x | x | x | x | x | x |
| Gender | x | x | x | x | x | x | x |
| Region | x | x | x | x | x | x | x |
| Household status | x | x | x | x | x | x | x |
| Occupation/Setting | x | x | x | x | x | x | x |

| **Cox proportional hazards models** | Cox model 1 | Cox model 2 | Cox model 3 | Cox model 4 |
|---|---|---|---|---|
| N (individuals in model) | 9,560 | 9,560 | 7,549 | 7,549 |
| Symptom data (Y/N) | N | N | N | N |
| **Covariates** | | | | |
| Month | | | | |
| Binary vaccination status | x | | x | |
| Vaccination status | | x | | |
| Time since infection | | | x | x |
| Vaccination:time since infection (interaction term) | | | x | x |
| Age group | s | s | s | s |
| Gender | x | x | x | x |
| Region | x | x | x | x |
| Household status | s | s | s | s |
| Occupation/Setting | s | s | s | s |

Table S3: Demographic characteristics of study participants.

| Characteristic | Overall | 2+ years | 1-2 years | 6-12 months | 0-6 months | No evidence of infection | Confirmed naïve |
|---|---|---|---|---|---|---|---|
| All | 9,560 | 1,105 | 1,153 | 2,378 | 2,140 | 2,100 | 773 |
| **Gender** | | | | | | | |
| Male | 1,524 (16%) | 205 (19%) | 180 (16%) | 382 (16%) | 346 (16%) | 298 (15%) | 113 (15%) |
| Female | 8,036 (84%) | 900 (81%) | 973 (84%) | 1,996 (84%) | 1,794 (84%) | 1,713 (85%) | 660 (85%) |
| **Age group** | | | | | | | |
| <25 | 71 (0.7%) | 5 (0.5%) | 15 (1.3%) | 24 (1.0%) | 16 (0.7%) | 9 (0.4%) | 2 (0.3%) |
| 25-34 | 828 (8.7%) | 62 (5.6%) | 95 (8.2%) | 243 (10%) | 222 (10%) | 145 (7.2%) | 61 (7.9%) |
| 35-44 | 2,096 (22%) | 211 (19%) | 244 (21%) | 691 (29%) | 447 (21%) | 376 (19%) | 127 (16%) |
| 45-54 | 3,702 (39%) | 434 (39%) | 456 (40%) | 881 (37%) | 824 (39%) | 788 (39%) | 319 (41%) |
| 55-64 | 2,637 (28%) | 371 (34%) | 317 (27%) | 501 (21%) | 580 (27%) | 625 (31%) | 243 (31%) |
| 65+ | 226 (2.4%) | 22 (2.0%) | 26 (2.3%) | 38 (1.6%) | 51 (2.4%) | 68 (3.4%) | 21 (2.7%) |
| **Ethnicity** | | | | | | | |
| White | 8,611 (90%) | 952 (86%) | 1,005 (87%) | 2,117 (89%) | 1,958 (91%) | 1,863 (93%) | 716 (93%) |
| Asian | 543 (5.7%) | 88 (8.0%) | 78 (6.8%) | 157 (6.6%) | 99 (4.6%) | 86 (4.3%) | 35 (4.5%) |
| Black | 179 (1.9%) | 35 (3.2%) | 42 (3.6%) | 35 (1.5%) | 29 (1.4%) | 29 (1.4%) | 9 (1.2%) |
| Mixed-race | 126 (1.3%) | 14 (1.3%) | 12 (1.0%) | 43 (1.8%) | 34 (1.6%) | 18 (0.9%) | 5 (0.6%) |
| Other | 101 (1.1%) | 16 (1.4%) | 16 (1.4%) | 26 (1.1%) | 20 (0.9%) | 15 (0.7%) | 8 (1.0%) |
| **Medical conditions** | | | | | | | |
| None | 7,038 (74%) | 805 (73%) | 855 (74%) | 1,759 (74%) | 1,582 (74%) | 1,458 (73%) | 579 (75%) |
| Immunosuppression | 247 (2.6%) | 37 (3.3%) | 21 (1.8%) | 60 (2.5%) | 56 (2.6%) | 51 (2.5%) | 22 (2.8%) |
| Chronic Respiratory conditions | 1,193 (12%) | 123 (11%) | 143 (12%) | 323 (14%) | 279 (13%) | 249 (12%) | 76 (9.8%) |
| Chronic Non-Respiratory conditions | 1,082 (11%) | 140 (13%) | 134 (12%) | 236 (9.9%) | 223 (10%) | 253 (13%) | 96 (12%) |
| **Staff type** | | | | | | | |
| Administrative/Executive (office based) | 1,613 (17%) | 184 (17%) | 185 (16%) | 389 (16%) | 338 (16%) | 377 (19%) | 140 (18%) |
| Doctor | 1,155 (12%) | 155 (14%) | 125 (11%) | 273 (11%) | 288 (13%) | 221 (11%) | 93 (12%) |
| Nursing | 3,178 (33%) | 362 (33%) | 388 (34%) | 814 (34%) | 699 (33%) | 647 (32%) | 268 (35%) |
| Healthcare Assistant | 584 (6.1%) | 82 (7.4%) | 78 (6.8%) | 144 (6.1%) | 115 (5.4%) | 115 (5.7%) | 50 (6.5%) |
| Midwife | 197 (2.1%) | 22 (2.0%) | 29 (2.5%) | 47 (2.0%) | 53 (2.5%) | 33 (1.6%) | 13 (1.7%) |
| Healthcare Scientist | 434 (4.5%) | 31 (2.8%) | 46 (4.0%) | 104 (4.4%) | 98 (4.6%) | 120 (6.0%) | 35 (4.5%) |
| Pharmacist | 268 (2.8%) | 20 (1.8%) | 34 (2.9%) | 69 (2.9%) | 59 (2.8%) | 52 (2.6%) | 34 (4.4%) |
| Physiotherapist/Occupational Therapist/SALT | 404 (4.2%) | 35 (3.2%) | 55 (4.8%) | 123 (5.2%) | 98 (4.6%) | 69 (3.4%) | 24 (3.1%) |
| Student (Medical/Nursing/Midwifery/Other) | 232 (2.4%) | 61 (5.5%) | 18 (1.6%) | 55 (2.3%) | 27 (1.3%) | 66 (3.3%) | 5 (0.6%) |
| Estates/Porters/Security | 185 (1.9%) | 14 (1.3%) | 36 (3.1%) | 41 (1.7%) | 55 (2.6%) | 32 (1.6%) | 7 (0.9%) |

**Table S3 continued from previous page**

| | | | | | | | |
|---|---|---|---|---|---|---|---|
| Other (non-patient facing) | 169 (1.8%) | 18 (1.6%) | 23 (2.0%) | 33 (1.4%) | 45 (2.1%) | 34 (1.7%) | 16 (2.1%) |
| Other (patient facing) | 1,141 (12%) | 121 (11%) | 136 (12%) | 286 (12%) | 265 (12%) | 245 (12%) | 88 (11%) |
| **Occupational setting** | | | | | | | |
| Outpatient | 3,841 (40%) | 386 (35%) | 450 (39%) | 961 (40%) | 995 (46%) | 675 (34%) | 374 (48%) |
| Ambulance/Emergency Department/Inpatient Wards | 396 (4.1%) | 39 (3.5%) | 56 (4.9%) | 116 (4.9%) | 98 (4.6%) | 47 (2.3%) | 40 (5.2%) |
| Intensive Care | 1,358 (14%) | 173 (16%) | 183 (16%) | 380 (16%) | 340 (16%) | 174 (8.7%) | 108 (14%) |
| Maternity/Labour Ward | 303 (3.2%) | 14 (1.3%) | 29 (2.5%) | 69 (2.9%) | 95 (4.4%) | 53 (2.6%) | 43 (5.6%) |
| Office | 829 (8.7%) | 108 (9.8%) | 90 (7.8%) | 206 (8.7%) | 108 (5.0%) | 286 (14%) | 31 (4.0%) |
| Other | 2,244 (23%) | 318 (29%) | 254 (22%) | 515 (22%) | 371 (17%) | 656 (33%) | 130 (17%) |
| Patient facing (non-clinical) | 502 (5.3%) | 56 (5.1%) | 78 (6.8%) | 114 (4.8%) | 120 (5.6%) | 91 (4.5%) | 43 (5.6%) |
| Theatres | 87 (0.9%) | 11 (1.0%) | 13 (1.1%) | 17 (0.7%) | 13 (0.6%) | 29 (1.4%) | 4 (0.5%) |
| **Patient contact** | | | | | | | |
| Yes | 7,992 (84%) | 943 (85%) | 983 (85%) | 2,008 (84%) | 1,796 (84%) | 1,631 (81%) | 631 (82%) |
| No | 1,568 (16%) | 162 (15%) | 170 (15%) | 370 (16%) | 344 (16%) | 380 (19%) | 142 (18%) |
| **Deprivation index** | | | | | | | |
| Most deprived (1) | 823 (9.1%) | 99 (9.1%) | 111 (10%) | 217 (9.8%) | 179 (9.0%) | 160 (8.3%) | 57 (7.8%) |
| Deprivation 2 | 1,528 (17%) | 183 (17%) | 210 (19%) | 408 (18%) | 312 (16%) | 314 (16%) | 101 (14%) |
| Deprivation 3 | 1,962 (22%) | 232 (21%) | 238 (22%) | 476 (21%) | 434 (22%) | 428 (22%) | 154 (21%) |
| Deprivation 4 | 2,121 (23%) | 260 (24%) | 259 (24%) | 498 (22%) | 474 (24%) | 459 (24%) | 171 (23%) |
| Least deprived (5) | 2,593 (29%) | 308 (28%) | 275 (25%) | 620 (28%) | 587 (30%) | 557 (29%) | 246 (34%) |
| **Region of residence** | | | | | | | |
| East Midlands | 517 (5.4%) | 78 (7.1%) | 72 (6.2%) | 124 (5.2%) | 35 (1.6%) | 182 (9.1%) | 26 (3.4%) |
| East of England | 1,176 (12%) | 155 (14%) | 172 (15%) | 287 (12%) | 296 (14%) | 116 (5.8%) | 150 (19%) |
| London | 1,199 (13%) | 202 (18%) | 174 (15%) | 326 (14%) | 235 (11%) | 212 (11%) | 50 (6.5%) |
| North East | 223 (2.3%) | 26 (2.4%) | 23 (2.0%) | 58 (2.4%) | 69 (3.2%) | 41 (2.0%) | 6 (0.8%) |
| North West | 794 (8.3%) | 117 (11%) | 89 (7.7%) | 208 (8.7%) | 171 (8.0%) | 143 (7.1%) | 66 (8.5%) |
| Northern Ireland | 499 (5.2%) | 22 (2.0%) | 60 (5.2%) | 151 (6.3%) | 146 (6.8%) | 77 (3.8%) | 43 (5.6%) |
| Scotland | 1,701 (18%) | 30 (2.7%) | 177 (15%) | 398 (17%) | 683 (32%) | 178 (8.9%) | 235 (30%) |
| South East | 947 (9.9%) | 133 (12%) | 88 (7.6%) | 226 (9.5%) | 110 (5.1%) | 340 (17%) | 50 (6.5%) |
| South West | 958 (10%) | 122 (11%) | 101 (8.8%) | 215 (9.0%) | 80 (3.7%) | 404 (20%) | 36 (4.7%) |
| Wales | 230 (2.4%) | 25 (2.3%) | 28 (2.4%) | 56 (2.4%) | 80 (3.7%) | 18 (0.9%) | 23 (3.0%) |
| West Midlands | 700 (7.3%) | 94 (8.5%) | 111 (9.6%) | 182 (7.7%) | 115 (5.4%) | 135 (6.7%) | 63 (8.2%) |
| Yorkshire and the Humber | 616 (6.4%) | 101 (9.1%) | 58 (5.0%) | 147 (6.2%) | 120 (5.6%) | 165 (8.2%) | 25 (3.2%) |
| **Household structure** | | | | | | | |
| Lives with others (no children) | 4,834 (51%) | 616 (56%) | 567 (49%) | 1,075 (45%) | 1,089 (51%) | 1,082 (54%) | 405 (52%) |
| Lives alone | 1,156 (12%) | 123 (11%) | 136 (12%) | 223 (9.4%) | 257 (12%) | 282 (14%) | 135 (17%) |

**Table S3 continued from previous page**

| | | | | | | | |
|---|---|---|---|---|---|---|---|
| Lives with others (with children) | 3,570 (37%) | 366 (33%) | 450 (39%) | 1,080 (45%) | 794 (37%) | 647 (32%) | 233 (30%) |

Table S4: Crude incidence calculations and protection from prior infection for selected covariates, by demographic characteristic.

| **Characteristic** | **Number of participants** | **Positive PCR tests** | **Exposure (person-days at risk)** | **Crude PCR positivity per 10,000 person-days (unadjusted) (95% CI)** | **Protection relative to baseline (fully adjusted) (95% CI)** |
|---|---|---|---|---|---|
| All | 9560 | 1298 | 1521928 | 8.53 (8.07, 9.01) | |
| **Age group** | | | | | |
| <25 | 71 | 2 | 9274 | 2.16 (0.26, 7.79) | 60.06% (-61.15, 90.1) |
| 25-34 | 828 | 131 | 118750 | 11.03 (9.22, 13.09) | -55.17% (-90.75, -26.23) |
| 35-44 | 2096 | 305 | 325934 | 9.36 (8.34, 10.47) | -15.73% (-35.22, 0.95) |
| 45-54 | 3702 | 498 | 594357 | 8.38 (7.66, 9.15) | Baseline |
| 55-64 | 2637 | 335 | 435942 | 7.68 (6.88, 8.55) | 7.84% (-7.84, 21.24) |
| 65+ | 226 | 27 | 37671 | 7.17 (4.72, 10.43) | 4.6% (-43.93, 36.77) |
| **Gender** | | | | | |
| Male | 1524 | 193 | 244177 | 7.9 (6.83, 9.1) | Baseline |
| Female | 8036 | 1105 | 1277751 | 8.65 (8.15, 9.17) | -9.83% (-29.54, 6.89) |
| **Ethnicity** | | | | | |
| White | 8611 | 1191 | 1378760 | 8.64 (8.15, 9.14) | |
| Asian | 543 | 64 | 81792 | 7.82 (6.03, 9.99) | |
| Black | 179 | 20 | 27264 | 7.34 (4.48, 11.33) | |
| Mixed-race | 126 | 12 | 19324 | 6.21 (3.21, 10.85) | |
| Other | 101 | 11 | 14788 | 7.44 (3.71, 13.31) | |
| **Medical conditions** | | | | | |
| None | 7038 | 955 | 1125891 | 8.48 (7.95, 9.04) | |
| Immunosuppression | 247 | 31 | 38272 | 8.1 (5.5, 11.5) | |
| Chronic Respiratory conditions | 1193 | 168 | 188356 | 8.92 (7.62, 10.37) | |
| Chronic Non-Respiratory conditions | 1082 | 144 | 169409 | 8.5 (7.17, 10.01) | |
| **Staff type** | | | | | |
| Administrative/Executive (office based) | 1613 | 197 | 258200 | 7.63 (6.6, 8.77) | |
| Doctor | 1155 | 156 | 185499 | 8.41 (7.14, 9.84) | |
| Nursing | 3178 | 437 | 499514 | 8.75 (7.95, 9.61) | |
| Healthcare Assistant | 584 | 95 | 92807 | 10.24 (8.28, 12.51) | |

**Table S4 continued from previous page**

| | | | | | |
|---|---|---|---|---|---|
| Midwife | 197 | 14 | 29522 | 4.74 (2.59, 7.96) | |
| Healthcare Scientist | 434 | 66 | 72365 | 9.12 (7.05, 11.6) | |
| Pharmacist | 268 | 51 | 43138 | 11.82 (8.8, 15.54) | |
| Physiotherapist/Occupational Therapist/SALT | 404 | 53 | 63614 | 8.33 (6.24, 10.9) | |
| Student (Medical/Nursing/Midwifery/Other) | 232 | 21 | 41009 | 5.12 (3.17, 7.83) | |
| Estates/Porters/Security | 185 | 23 | 29218 | 7.87 (4.99, 11.81) | |
| Other (non-patient facing) | 169 | 27 | 29060 | 9.29 (6.12, 13.52) | |
| Other (patient facing) | 1141 | 158 | 177982 | 8.88 (7.55, 10.37) | |
| **Setting** | | | | | |
| Outpatient | 3841 | 568 | 600914 | 9.45 (8.69, 10.26) | Baseline |
| Ambulance/Emergency Department/Inpatient Wards | 396 | 65 | 58022 | 11.2 (8.65, 14.28) | -9.56% (-44.84, 17.13) |
| Intensive Care | 1358 | 209 | 200329 | 10.43 (9.07, 11.95) | -14.19% (-35.1, 3.48) |
| Maternity/Labour Ward | 303 | 47 | 50155 | 9.37 (6.89, 12.46) | 10.06% (-22.79, 34.12) |
| Office | 829 | 81 | 145247 | 5.58 (4.43, 6.93) | 16.61% (-7.57, 35.36) |
| Other | 2244 | 254 | 376276 | 6.75 (5.95, 7.63) | 7.11% (-9.71, 21.36) |
| Patient facing (non-clinical) | 502 | 58 | 76398 | 7.59 (5.76, 9.81) | 1.92% (-30.01, 26.01) |
| Theatres | 87 | 16 | 14587 | 10.97 (6.27, 17.81) | -83.43% (-204.47, -10.51) |
| **Patient contact** | | | | | |
| Yes | 7992 | 1086 | 1266739 | 8.57 (8.07, 9.1) | |
| No | 1568 | 212 | 255189 | 8.31 (7.23, 9.5) | |
| **Deprivation index** | | | | | |
| Most deprived (1) | 823 | 112 | 129010 | 8.68 (7.15, 10.45) | |
| Deprivation 2 | 1528 | 194 | 241221 | 8.04 (6.95, 9.26) | |
| Deprivation 3 | 1962 | 257 | 310875 | 8.27 (7.29, 9.34) | |
| Deprivation 4 | 2121 | 282 | 333791 | 8.45 (7.49, 9.49) | |
| Least deprived (5) | 2593 | 361 | 421315 | 8.57 (7.71, 9.5) | |
| **Region of residence** | | | | | |
| East Midlands | 517 | 56 | 89779 | 6.24 (4.71, 8.1) | Baseline |
| East of England | 1176 | 160 | 195308 | 8.19 (6.97, 9.56) | -1.24% (-41.2, 27.41) |
| London | 1199 | 102 | 182727 | 5.58 (4.55, 6.78) | 33.66% (4.85, 53.75) |
| North East | 223 | 31 | 29820 | 10.4 (7.06, 14.76) | -0.77% (-61.97, 37.3) |
| North West | 794 | 113 | 111247 | 10.16 (8.37, 12.21) | -38.68% (-96.56, 2.16) |
| Northern Ireland | 499 | 89 | 80473 | 11.06 (8.88, 13.61) | -13.44% (-63.67, 21.38) |
| Scotland | 1701 | 330 | 275189 | 11.99 (10.73, 13.36) | -18.65% (-62.68, 13.47) |
| South East | 947 | 98 | 156100 | 6.28 (5.1, 7.65) | -1.04% (-44.54, 29.36) |
| South West | 958 | 97 | 168695 | 5.75 (4.66, 7.01) | -9.22% (-56.33, 23.69) |

**Table S4 continued from previous page**

| | | | | | |
|---|---|---|---|---|---|
| Wales | 230 | 58 | 39407 | 14.72 (11.18, 19.03) | -47.2% (-120.42, 1.69) |
| West Midlands | 700 | 78 | 98077 | 7.95 (6.29, 9.93) | -1.9% (-48.78, 30.21) |
| Yorkshire and the Humber | 616 | 86 | 95106 | 9.04 (7.23, 11.17) | -2.82% (-49.45, 29.26) |
| **Household structure** | | | | | |
| Lives with others (no children) | 4834 | 632 | 777602 | 8.13 (7.51, 8.79) | Baseline |
| Lives alone | 1156 | 161 | 187571 | 8.58 (7.31, 10.02) | 1.2% (-18.94, 17.93) |
| Lives with others (with children) | 3570 | 505 | 556755 | 9.07 (8.3, 9.9) | -4.38% (-20.03, 9.23) |
| **Month of study** | | | | | |
| Sep 2022 | 9559 | 105 | 176165 | 5.96 (4.87, 7.22) | |
| Oct 2022 | 9025 | 181 | 271290 | 6.67 (5.74, 7.72) | |
| Nov 2022 | 8515 | 143 | 249090 | 5.74 (4.84, 6.76) | |
| Dec 2022 | 8096 | 292 | 243641 | 11.98 (10.65, 13.44) | |
| Jan 2023 | 7669 | 193 | 227909 | 8.47 (7.32, 9.75) | |
| Feb 2023 | 6943 | 182 | 181605 | 10.02 (8.62, 11.59) | |
| Mar 2023 | 5772 | 202 | 172228 | 11.73 (10.17, 13.46) | |

Table S5: Crude incidence calculations and estimates of relative vaccine effectiveness by vaccination status and time since previous infection.

| Time since previous infection | Vaccination status | Number of participants | Positive PCR tests | Exposure (person-days at risk) | Crude PCR positivity per 10,000 person-days (unadjusted) (95% CI) | VE relative to baseline (fully adjusted) (95% CI) |
|---|---|---|---|---|---|---|
| Confirmed naïve | Waned third dose | 755 | 51 | 45366 | 11.24 (8.37, 14.78) | Baseline |
| | Fourth dose | 527 | 122 | 69799 | 17.48 (14.52, 20.87) | -30.87% (-84.79, 7.31) |
| 2+ years | Waned third dose | 1345 | 77 | 82640 | 9.32 (7.35, 11.65) | Baseline |
| | Fourth dose | 1183 | 119 | 140143 | 8.49 (7.03, 10.16) | 32.87% (8.51, 52.20) |
| 1-2 years | Waned third dose | 1679 | 83 | 80251 | 10.34 (8.24, 12.82) | Baseline |
| | Fourth dose | 1845 | 126 | 115464 | 10.91 (9.09, 12.99) | -1.67% (-40.95, 26.66) |
| 6-12 months | Waned third dose | 3136 | 159 | 174120 | 9.13 (7.77, 10.67) | Baseline |
| | Fourth dose | 2659 | 188 | 252695 | 7.44 (6.41, 8.58) | 23.14% (3.07, 39.06) |
| 0-6 months | Waned third dose | 2851 | 45 | 121019 | 3.72 (2.71, 4.98) | Baseline |
| | Fourth dose | 1779 | 38 | 133847 | 2.84 (2.01, 3.9) | 39.58% (7.99, 60.32) |

Table S6: Estimated time spent in PCR positive state, averaged across the study population, and empirical median duration of PCR positivity by vaccination status and time since previous infection.

| | **Empirical median duration of PCR positivity (initial PCR positive to subsequent PCR negative) [interquartile range]** | **Estimated days spent in PCR positive state (95% CI)** |
|---|---|---|
| Whole population | 15 days [12, 27] | 7.51 days (6.94, 8.13) |
| **Vaccination status** | | |
| Waned third dose | 15 days [13, 28] | 8.50 days (6.79, 10.64) |
| Fourth dose | 14 days [12, 26] | 6.90 days (5.87, 8.11) |
| **Time since previous infection** | | |
| Confirmed naive | 16 days [13, 28] | 9.51 days (7.08, 12.78) |
| 2+ years | 14 days [12, 27] | 9.16 days (7.16, 11.72) |
| 1-2 years | 14 days [12, 21] | 7.44 days (5.86, 9,44) |
| 6-12 months | 14 days [12, 23] | 6.56 days (5.63, 7.63) |
| 0-6 months | 16 days [14, 28] | 7.25 days (6.32, 8.31) |
| **COVID symptoms** | | |
| COVID symptoms | 15 days [13, 27] | 8.09 days (7.36, 8.90) |
| Non-COVID symptoms or asymptomatic | 14 days [12, 23] | 4.70 days (4.04, 5.47) |

[11] V. Hall, S. Foulkes, F. Insalata, P. Kirwan, A. Saei, A. Atti, E. Wellington, J. Khawam, K. Munro, M. Cole, C. Tranquillini, A. Taylor-Kerr, N. Hettiarachchi, D. Calbraith, N. Sajedi, I. Milligan, Y. Themistocleous, D. Corrigan, L. Cromey, L. Price, S. Stewart, E. de Lacy, C. Norman, E. Linley, A. D. Otter, A. Semper, J. Hewson, S. D'Arcangelo, M. Chand, C. S. Brown, T. Brooks, J. Islam, A. Charlett, S. Hopkins, and SIREN

Study Group. “Protection against SARS-CoV-2 after Covid-19 Vaccination and Previous Infection”. en. In: *N. Engl. J. Med.* (Feb. 2022). URL: http://dx.doi.org/10.1056/NEJMoa2118691.

[12] J. M. Gran, S. A. Lie, I. Øyeflaten, Ø. Borgan, and O. O. Aalen. “Causal inference in multi-state models–sickness absence and work for 1145 participants after work rehabilitation”. en. In: *BMC Public Health* 15.1 (Oct. 2015), pp. 1–16. URL: https://bmcpublichealth.biomedcentral.com/articles/10.1186/s12889-015-2408-8.
