## Supplementary material for "Protection of second booster vaccinations and prior infection against SARS-CoV-2 in the UK SIREN healthcare worker cohort": SIREN Study Group

| Organisation Name | First Name/Initial | Surname |
|---|---|---|
| **Participating NHS Sites** | | |
| HYWEL DDA UNIVERSITY LHB | Tracy | Lewis |
| SWANSEA BAY UNIVERSITY LHB | Steve | Bain |
| SWANSEA BAY UNIVERSITY LHB | Rebeccah | Thomas |
| CWM TAF MORGANNWG UNIVERSITY LHB | John | Geen |
| CWM TAF MORGANNWG UNIVERSITY LHB | Carla | Pothecary |
| ANEURIN BEVAN UNIVERSITY LHB | Sean | Cutler |
| ANEURIN BEVAN UNIVERSITY LHB | John | Northfield |
| SOLENT NHS TRUST | Cathy | Price |
| SHROPSHIRE COMMUNITY HEALTH NHS TRUST | Johanne | Tomlinson |
| ISLE OF WIGHT NHS TRUST | Sarah | Knight |
| ISLE OF WIGHT NHS TRUST | Emily | Macnaughton |
| LONDON NORTH WEST UNIVERSITY HEALTHCARE NHS TRUST | Ekaterina | Watson |
| UNIVERSITY HOSPITALS BRISTOL AND WESTON NHS FOUNDATION TRUST | Rajeka | Lazarus |
| UNIVERSITY HOSPITALS BRISTOL AND WESTON NHS FOUNDATION TRUST | Aaran | Sinclair |
| SOUTHEND UNIVERSITY HOSPITAL NHS FOUNDATION TRUST | Joanne | Galliford |
| SOUTHEND UNIVERSITY HOSPITAL NHS FOUNDATION TRUST | Bridgett | Masunda |
| ROYAL FREE LONDON NHS FOUNDATION TRUST | Tabitha | Mahungu |
| ROYAL FREE LONDON NHS FOUNDATION TRUST | Alison | Rodger |
| ROYAL NATIONAL ORTHOPAEDIC HOSPITAL NHS TRUST | Esther | Hanison |
| ROYAL NATIONAL ORTHOPAEDIC HOSPITAL NHS TRUST | Simon | Warren |
| NORTH MIDDLESEX UNIVERSITY HOSPITAL NHS TRUST | Swati | Jain |
| NORTH MIDDLESEX UNIVERSITY HOSPITAL NHS TRUST | Mariyam | Mirfenderesky |
| THE HILLINGDON HOSPITALS NHS FOUNDATION TRUST | Natasha | Mahabir |
| ST HELENS AND KNOWSLEY TEACHING HOSPITALS NHS TRUST | Rowan | Pritchard-Jones |
| ST HELENS AND KNOWSLEY TEACHING HOSPITALS NHS TRUST | Diane | Wycherley |
| MID CHESHIRE HOSPITALS NHS FOUNDATION TRUST | Claire | Gabriel |
| MID CHESHIRE HOSPITALS NHS FOUNDATION TRUST | Elijah | Matovu |
| BEDFORDSHIRE HOSPITALS NHS FOUNDATION TRUST | Philippa | Bakker |
| BEDFORDSHIRE HOSPITALS NHS FOUNDATION TRUST | Simantee | Guha |
| SHEFFIELD CHILDREN'S NHS FOUNDATION TRUST | S | Gormley |
| SHEFFIELD CHILDREN'S NHS FOUNDATION TRUST | James | Pethick |
| BASILDON AND THURROCK UNIVERSITY HOSPITALS NHS FOUNDATION TRUST | Georgina | Butt |
| BASILDON AND THURROCK UNIVERSITY HOSPITALS NHS FOUNDATION TRUST | Stacey | Pepper |
| EAST SUFFOLK AND NORTH ESSEX NHS FOUNDATION TRUST | Luke | Bedford |
| EAST SUFFOLK AND NORTH ESSEX NHS FOUNDATION TRUST | Paul | Ridley |
| FRIMLEY HEALTH NHS FOUNDATION TRUST | Jane | Democratis |

| | | |
|---|---|---|
| FRIMLEY HEALTH NHS FOUNDATION TRUST | Manjula | Meda |
| LIVERPOOL UNIVERSITY HOSPITALS NHS FOUNDATION TRUST | Anu | Chawla |
| LIVERPOOL UNIVERSITY HOSPITALS NHS FOUNDATION TRUST | Fran | Westwell |
| THE CLATTERBRIDGE CANCER CENTRE NHS FOUNDATION TRUST | Nagesh | Kalakonda |
| THE CLATTERBRIDGE CANCER CENTRE NHS FOUNDATION TRUST | Sheena | Khanduri |
| ROYAL PAPWORTH HOSPITAL NHS FOUNDATION TRUST | Allison | Doel |
| ROYAL PAPWORTH HOSPITAL NHS FOUNDATION TRUST | Sumita | Pai |
| JAMES PAGET UNIVERSITY HOSPITALS NHS FOUNDATION TRUST | Christian | Hacon |
| JAMES PAGET UNIVERSITY HOSPITALS NHS FOUNDATION TRUST | Davis | Nwaka |
| WEST SUFFOLK NHS FOUNDATION TRUST | Veronica | Mendez Moro |
| WEST SUFFOLK NHS FOUNDATION TRUST | A | Moody |
| ROYAL DEVON AND EXETER NHS FOUNDATION TRUST | Cressida | Auckland |
| ROYAL DEVON AND EXETER NHS FOUNDATION TRUST | Stephanie | Prince |
| SHEFFIELD TEACHING HOSPITALS NHS FOUNDATION TRUST | Thushan | de Silva |
| SHEFFIELD TEACHING HOSPITALS NHS FOUNDATION TRUST | Helen | Shulver |
| LEWISHAM AND GREENWICH NHS TRUST | A | Shah |
| CROYDON HEALTH SERVICES NHS TRUST | C | Jones |
| CROYDON HEALTH SERVICES NHS TRUST | Banerjee | Subhro-Osuji |
| ST GEORGE'S UNIVERSITY HOSPITALS NHS FOUNDATION TRUST | Angela | Houston |
| ST GEORGE'S UNIVERSITY HOSPITALS NHS FOUNDATION TRUST | Tim | Planche |
| UNIVERSITY HOSPITALS OF NORTH MIDLANDS NHS TRUST | Martin | Booth |
| UNIVERSITY HOSPITALS OF NORTH MIDLANDS NHS TRUST | Christopher | Duff |
| KING'S COLLEGE HOSPITAL NHS FOUNDATION TRUST | Jonnie | Aeron-Thomas |
| KING'S COLLEGE HOSPITAL NHS FOUNDATION TRUST | Ray | Chaudhuri |
| UNIVERSITY HOSPITALS PLYMOUTH NHS TRUST | David | Hilton |
| UNIVERSITY HOSPITALS PLYMOUTH NHS TRUST | Hannah | Jory |
| WHITTINGTON HEALTH NHS TRUST | Zehra'a | Al-Khafaji |
| WHITTINGTON HEALTH NHS TRUST | Philippa | Kemsley |
| THE ROBERT JONES AND AGNES HUNT ORTHOPAEDIC HOSPITAL NHS FOUNDATION TRUST | Ruth | Longfellow |
| GEORGE ELIOT HOSPITAL NHS TRUST | David | Boss |
| GEORGE ELIOT HOSPITAL NHS TRUST | Simon | Brake |
| NORFOLK AND NORWICH UNIVERSITY HOSPITALS NHS FOUNDATION TRUST | Louise | Coke |
| NORFOLK AND NORWICH UNIVERSITY HOSPITALS NHS FOUNDATION TRUST | Ngozi | Elumogo |
| BOLTON NHS FOUNDATION TRUST | Scott | Latham |
| BOLTON NHS FOUNDATION TRUST | Chinari | Subudhi |
| HAMPSHIRE HOSPITALS NHS FOUNDATION TRUST | Ina | Hoad |
| HAMPSHIRE HOSPITALS NHS FOUNDATION TRUST | Claire | Thomas |

| | | |
|---|---|---|
| DARTFORD AND GRAVESHAM NHS TRUST | Nihil | Chitalia |
| DARTFORD AND GRAVESHAM NHS TRUST | Tracy | Edmunds |
| THE DUDLEY GROUP NHS FOUNDATION TRUST | Helen | Ashby |
| NORTH CUMBRIA INTEGRATED CARE NHS FOUNDATION TRUST | John | Elliott |
| NORTH CUMBRIA INTEGRATED CARE NHS FOUNDATION TRUST | Beverley | Wilkinson |
| SALISBURY NHS FOUNDATION TRUST | Abby | Rand |
| SALISBURY NHS FOUNDATION TRUST | Catherine | Thompson |
| DONCASTER AND BASSETLAW TEACHING HOSPITALS NHS FOUNDATION TRUST | K | Agwuh |
| DONCASTER AND BASSETLAW TEACHING HOSPITALS NHS FOUNDATION TRUST | Anna | Grice |
| LINCOLNSHIRE PARTNERSHIP NHS FOUNDATION TRUST | Kelly | Moran |
| LINCOLNSHIRE PARTNERSHIP NHS FOUNDATION TRUST | Vijayendra | Waykar |
| MID ESSEX HOSPITAL SERVICES NHS TRUST | Yvonne | Lester |
| MID ESSEX HOSPITAL SERVICES NHS TRUST | Lauren | Sach |
| THE PRINCESS ALEXANDRA HOSPITAL NHS TRUST | Kathryn | Court |
| THE PRINCESS ALEXANDRA HOSPITAL NHS TRUST | Nikki | White |
| LEEDS TEACHING HOSPITALS NHS TRUST | Clair | Favager |
| LEEDS TEACHING HOSPITALS NHS TRUST | Kyra | Holliday |
| THE NEWCASTLE UPON TYNE HOSPITALS NHS FOUNDATION TRUST | Jayne | Harwood |
| THE NEWCASTLE UPON TYNE HOSPITALS NHS FOUNDATION TRUST | Brendan | Payne |
| UNIVERSITY HOSPITALS OF MORECAMBE BAY NHS FOUNDATION TRUST | Karen | Burns |
| UNIVERSITY HOSPITALS OF MORECAMBE BAY NHS FOUNDATION TRUST | Lynda | Fothergill |
| CENTRAL AND NORTH WEST LONDON NHS FOUNDATION TRUST | Alejandro | Arenas-Pinto |
| CENTRAL AND NORTH WEST LONDON NHS FOUNDATION TRUST | Abigail | Severn |
| SOUTHPORT AND ORMSKIRK HOSPITAL NHS TRUST | Kerryanne | Brown |
| SOUTHPORT AND ORMSKIRK HOSPITAL NHS TRUST | Katherine | Gray |
| SOUTHERN HEALTH NHS FOUNDATION TRUST | Jane | Dare |
| SOUTHERN HEALTH NHS FOUNDATION TRUST | Qi | Zheng |
| LANCASHIRE & SOUTH CUMBRIA NHS FOUNDATION TRUST | Kathryn | Hollinshead |
| LANCASHIRE & SOUTH CUMBRIA NHS FOUNDATION TRUST | Robert | Shorten |
| UNITED LINCOLNSHIRE HOSPITALS NHS TRUST | Alun | Roebuck |
| UNIVERSITY HOSPITALS OF LEICESTER NHS TRUST | Christopher | Holmes |
| UNIVERSITY HOSPITALS OF LEICESTER NHS TRUST | Martin | Wiselka |
| STOCKPORT NHS FOUNDATION TRUST | Barzo | Faris |
| STOCKPORT NHS FOUNDATION TRUST | Liane | Marsh |
| DEVON PARTNERSHIP NHS TRUST | Cressida | Auckland |
| DEVON PARTNERSHIP NHS TRUST | Clare | McAdam |
| WARRINGTON AND HALTON TEACHING HOSPITALS NHS FOUNDATION TRUST | Lisa | Ditchfield |

| | | |
|---|---|---|
| WARRINGTON AND HALTON TEACHING HOSPITALS NHS FOUNDATION TRUST | Zaman | Qazzafi |
| CALDERDALE AND HUDDERSFIELD NHS FOUNDATION TRUST | G | Boyd |
| CALDERDALE AND HUDDERSFIELD NHS FOUNDATION TRUST | N | Wong |
| NOTTINGHAM UNIVERSITY HOSPITALS NHS TRUST | Sarah | Brand |
| NOTTINGHAM UNIVERSITY HOSPITALS NHS TRUST | Jack | Squires |
| MID YORKSHIRE HOSPITALS NHS TRUST | John | Ashcroft |
| MID YORKSHIRE HOSPITALS NHS TRUST | Ismaelette | Del Rosario |
| BLACKPOOL TEACHING HOSPITALS NHS FOUNDATION TRUST | Joanne | Howard |
| BLACKPOOL TEACHING HOSPITALS NHS FOUNDATION TRUST | Emma | Ward |
| DERBYSHIRE HEALTHCARE NHS FOUNDATION TRUST | Gemma | Harrison |
| DERBYSHIRE HEALTHCARE NHS FOUNDATION TRUST | Joely | Morgan |
| LANCASHIRE TEACHING HOSPITALS NHS FOUNDATION TRUST | Claire | Corless |
| LANCASHIRE TEACHING HOSPITALS NHS FOUNDATION TRUST | Robert | Shorten |
| BUCKINGHAMSHIRE HEALTHCARE NHS TRUST | Ruth | Penn |
| BUCKINGHAMSHIRE HEALTHCARE NHS TRUST | Nick | Wong |
| BIRMINGHAM AND SOLIHULL MENTAL HEALTH NHS FOUNDATION TRUST | Manny | Bagary |
| BIRMINGHAM AND SOLIHULL MENTAL HEALTH NHS FOUNDATION TRUST | Nadezda | Starkova |
| SHREWSBURY AND TELFORD HOSPITAL NHS TRUST | Mandy | Beekes |
| SHREWSBURY AND TELFORD HOSPITAL NHS TRUST | Mandy | Carnahan |
| HOUNSLOW AND RICHMOND COMMUNITY HEALTHCARE NHS TRUST | Shivani | Khan |
| HOUNSLOW AND RICHMOND COMMUNITY HEALTHCARE NHS TRUST | Shekoo | Mackay |
| IMPERIAL COLLEGE HEALTHCARE NHS TRUST | Keneisha | Lewis |
| IMPERIAL COLLEGE HEALTHCARE NHS TRUST | Graham | Pickard |
| NHS BORDERS | Joy | Dawson |
| NHS BORDERS | Lauren | Finlayson |
| NHS FORTH VALLEY | Euan | Cameron |
| NHS FORTH VALLEY | Anne | Todd |
| NHS GRAMPIAN | Sebastien | Fagegaltier |
| NHS GRAMPIAN | Sally | Mavin |
| NHS HIGHLAND | Alexandra | Cochrane |
| NHS HIGHLAND | Andrew | Gibson |
| NHS LOTHIAN | Sam | Donaldson |
| NHS LOTHIAN | Kate | Templeton |
| NHS WESTERN ISLES | Martin | Malcolm |
| NHS WESTERN ISLES | Beth | Smith |
| NHS FIFE | Devesh | Dhasmana |
| NHS FIFE | Susan | Fowler |
| NHS GREATER GLASGOW AND CLYDE | Antonia | Ho |
| NHS GREATER GLASGOW AND CLYDE | Michael | Murphy |
| NHS LANARKSHIRE | Claire | Beith |

| | | |
|---|---|---|
| NHS LANARKSHIRE | Manish | Patel |
| GOLDEN JUBILEE NATIONAL HOSPITAL | Elizabeth | Boyd |
| GOLDEN JUBILEE NATIONAL HOSPITAL | Val | Irvine |
| UNIVERSITY OF ST ANDREWS | David | Crossman |
| BLACK COUNTRY HEALTHCARE NHS FOUNDATION TRUST | Alison | Grant |
| BLACK COUNTRY HEALTHCARE NHS FOUNDATION TRUST | Rebecca | Temple-Purcell |
| BELFAST HEALTH & SOCIAL CARE TRUST | Clodagh | Loughrey |
| NORTHERN HEALTH & SOCIAL CARE TRUST | Elinor | Hanna |
| NORTHERN HEALTH & SOCIAL CARE TRUST | Frances | Johnston |
| SOUTHERN HEALTH & SOCIAL CARE TRUST | Angel | Boulos |
| SOUTHERN HEALTH & SOCIAL CARE TRUST | Fiona | Thompson |
| SOUTH EASTERN HEALTH & SOCIAL CARE | Yuri | Protaschik |
| SOUTH EASTERN HEALTH & SOCIAL CARE | Susan | Regan |
| WESTERN HEALTH & SOCIAL CARE TRUST | Tracy | Donaghy |
| WESTERN HEALTH & SOCIAL CARE TRUST | Maurice | O'Kane |
| **SIREN Study team** | | |
| UK HEALTH SECURITY AGENCY | Omolola | Akinbami |
| UK HEALTH SECURITY AGENCY | Paola | Barbero |
| UK HEALTH SECURITY AGENCY | Tim | Brooks |
| UK HEALTH SECURITY AGENCY | Meera | Chand |
| UK HEALTH SECURITY AGENCY | Andre | Charlett |
| UK HEALTH SECURITY AGENCY | Michelle | Cole |
| UK HEALTH SECURITY AGENCY | Ferdinando | Insalata |
| UK HEALTH SECURITY AGENCY | Palak | Joshi |
| UK HEALTH SECURITY AGENCY | Anne-Marie | O’Connell |
| UK HEALTH SECURITY AGENCY | Mary | Ramsay |
| UK HEALTH SECURITY AGENCY | Ayoub | Saei |
| UK HEALTH SECURITY AGENCY | Maria | Zambon |
| UK HEALTH SECURITY AGENCY | Ezra | Linley |
| UK HEALTH SECURITY AGENCY | Simon | Tonge |
| UK HEALTH SECURITY AGENCY | Enemona | Adaji |
| UK HEALTH SECURITY AGENCY | Omoyeni | Adebiyi |
| UK HEALTH SECURITY AGENCY | Nick | Andrews |
| UK HEALTH SECURITY AGENCY | Ana | Atti |
| UK HEALTH SECURITY AGENCY | Jonathan | Broad |
| UK HEALTH SECURITY AGENCY | Colin | Brown |
| UK HEALTH SECURITY AGENCY | Joanna | Conneely |
| UK HEALTH SECURITY AGENCY | Paul | Conneely |
| UK HEALTH SECURITY AGENCY | Angela | Dunne |
| UK HEALTH SECURITY AGENCY | Simone | Dyer |
| UK HEALTH SECURITY AGENCY | Hannah | Emmett |
| UK HEALTH SECURITY AGENCY | Sarah | Foulkes |
| UK HEALTH SECURITY AGENCY | Victoria | Hall |
| UK HEALTH SECURITY AGENCY | Nipunadi | Hettiarachchi |
| UK HEALTH SECURITY AGENCY | Susan | Hopkins |
| UK HEALTH SECURITY AGENCY | Anna | Howells |

| | | |
|---|---|---|
| UK HEALTH SECURITY AGENCY | Jasmin | Islam |
| UK HEALTH SECURITY AGENCY | Nishanthan | Kapirial |
| UK HEALTH SECURITY AGENCY | Jameel | Khawam |
| UK HEALTH SECURITY AGENCY | Edward | Monk |
| UK HEALTH SECURITY AGENCY | Katie | Munro |
| UK HEALTH SECURITY AGENCY | Naomi | Platt |
| UK HEALTH SECURITY AGENCY | Sophie | Russell |
| UK HEALTH SECURITY AGENCY | Dominic | Sparkes |
| UK HEALTH SECURITY AGENCY | Andrew | Taylor-Kerr |
| UK HEALTH SECURITY AGENCY | Jean | Timeyin |
| UK HEALTH SECURITY AGENCY | Silvia | D'Arcangelo |
| UK HEALTH SECURITY AGENCY | Ashley | Otter |
| UK HEALTH SECURITY AGENCY | Cathy | Rowe |
| UK HEALTH SECURITY AGENCY | Amanda | Semper |
| UK HEALTH SECURITY AGENCY | Eileen | Gallagher |
| UK HEALTH SECURITY AGENCY | Robert | Kyffin |
| PUBLIC HEALTH AGENCY NORTHERN IRELAND | Dianne | Corrigan |
| PUBLIC HEALTH AGENCY NORTHERN IRELAND | Lisa | Cromey |
| GLASGOW CALEDONIAN UNIVERSITY | Desmond | Areghan |
| PUBLIC HEALTH SCOTLAND | Jennifer | Bishop |
| GLASGOW CALEDONIAN UNIVERSITY | Melanie | Dembinsky |
| PUBLIC HEALTH SCOTLAND | Laura | Dobbie |
| PUBLIC HEALTH SCOTLAND | Josie | Evans |
| PUBLIC HEALTH SCOTLAND | David | Goldberg |
| GLASGOW CALEDONIAN UNIVERSITY & PUBLIC HEALTH SCOTLAND | Lynne | Haahr |
| GLASGOW CALEDONIAN UNIVERSITY | Annelysse | Jorgenson |
| GLASGOW CALEDONIAN UNIVERSITY | Ayodeji | Matuluko |
| PUBLIC HEALTH SCOTLAND | Laura | Naismith |
| GLASGOW CALEDONIAN UNIVERSITY & PUBLIC HEALTH SCOTLAND | Desy | Nuryunarsih |
| GLASGOW CALEDONIAN UNIVERSITY | Alexander | Olaoye |
| PUBLIC HEALTH SCOTLAND | Caitlin | Plank |
| GLASGOW CALEDONIAN UNIVERSITY & PUBLIC HEALTH SCOTLAND | Lesley | Price |
| GLASGOW CALEDONIAN UNIVERSITY & PUBLIC HEALTH SCOTLAND | Nicole | Sergenson |
| GLASGOW CALEDONIAN UNIVERSITY & PUBLIC HEALTH SCOTLAND | Sally | Stewart |
| PUBLIC HEALTH SCOTLAND | Andrew | Telfer |
| PUBLIC HEALTH SCOTLAND | Jennifer | Weir |
| PUBLIC HEALTH WALES | Ellen | De Lacy |
| HEALTH AND CARE RESEARCH WALES | Yvette | Ellis |
| PUBLIC HEALTH WALES | Susannah | Froude |
| HEALTH AND CARE RESEARCH WALES | Chris | Norman |
| PUBLIC HEALTH WALES | Guy | Stevens |
| PUBLIC HEALTH WALES | Linda | Tyson |
| MRC BIOSTATISTICS UNIT, UNIVERSITY OF CAMBRIDGE | Peter | Kirwan |
| MRC BIOSTATISTICS UNIT, UNIVERSITY OF CAMBRIDGE | Christopher | Jackson |

| | | |
|---|---|---|
| MRC BIOSTATISTICS UNIT, UNIVERSITY OF CAMBRIDGE | Anne | Presanis |
| MRC BIOSTATISTICS UNIT, UNIVERSITY OF CAMBRIDGE | Daniela | De Angelis |
| **SIREN Consortium** | | |
| PROTECTIVE IMMUNITY FROM T CELLS TO COVID-19 IN HEALTH WORKERS (PITCH), UNIVERSITY OF OXFORD | Susanna | Dunachie |
| PROTECTIVE IMMUNITY FROM T CELLS TO COVID-19 IN HEALTH WORKERS (PITCH), UNIVERSITY OF OXFORD | Paul | Klenerman |
| PROTECTIVE IMMUNITY FROM T CELLS TO COVID-19 IN HEALTH WORKERS (PITCH), UNIVERSITY OF NEWCASTLE | Chris | Duncan |
| PROTECTIVE IMMUNITY FROM T CELLS TO COVID-19 IN HEALTH WORKERS (PITCH), UNIVERSITY OF NEWCASTLE | Rebecca | Payne |
| PROTECTIVE IMMUNITY FROM T CELLS TO COVID-19 IN HEALTH WORKERS (PITCH), UNIVERSITY OF LIVERPOOL | Lance | Turtle |
| PROTECTIVE IMMUNITY FROM T CELLS TO COVID-19 IN HEALTH WORKERS (PITCH), UNIVERSITY OF BIRMINGHAM | Alex | Richter |
| PROTECTIVE IMMUNITY FROM T CELLS TO COVID-19 IN HEALTH WORKERS (PITCH), UNIVERSITY OF SHEFFIELD | Thushan | De Silva |
| PROTECTIVE IMMUNITY FROM T CELLS TO COVID-19 IN HEALTH WORKERS (PITCH), UNIVERSITY OF OXFORD | Eleanor | Barnes |
| PROTECTIVE IMMUNITY FROM T CELLS TO COVID-19 IN HEALTH WORKERS (PITCH), UNIVERSITY OF NEWCASTLE | Daniel | Wootton |
| VACCINE IMMUNITY, BREAKTHROUGH & REINFECTION - ANTIBODIES & T-CELLS (VIBRANT) STUDY, UNIVERSITY OF BIRMINGHAM | Oliver | Galgut |
| THE HUMORAL IMMUNE CORRELATES FOR COVID-19 (HICC) CONSORTIUM, UNIVERSITY OF CAMBRIDGE | Jonathan | Heeney |
| THE HUMORAL IMMUNE CORRELATES FOR COVID-19 (HICC) CONSORTIUM, UNIVERSITY OF CAMBRIDGE | Helen | Baxendale |
| THE HUMORAL IMMUNE CORRELATES FOR COVID-19 (HICC) CONSORTIUM, UNIVERSITY OF CAMBRIDGE | Javier | Castillo-Olivares |
| THE FRANCIS CRICK INSTITUTE | Rupert | Beale |
| THE FRANCIS CRICK INSTITUTE | Edward | Carr |
| GENOTYPE2PHENOTYPE (G2P) IMPERIAL COLLEGE LONDON | Wendy | Barclay |
| GENOTYPE2PHENOTYPE (G2P) IMPERIAL COLLEGE LONDON | Maya | Moshe |
| GENOTYPE2PHENOTYPE (G2P) UNIVERSITY OF GLASGOW | Massimo | Palmarini |
| GENOTYPE2PHENOTYPE (G2P) UNIVERSITY OF GLASGOW | Brian | Willett |
| GENOMICC, UNIVERSITY OF EDINBURGH | John Kenneth | Baillie |
| BRITISH SOCIETY FOR IMMUNOLOGY | Jennie | Evans |
| BRITISH SOCIETY FOR IMMUNOLOGY | Erika | Aquino |
